# A Multidimensional Immune Signature Predicts Susceptibility to Omicron Infection in Vaccinated Individuals

**DOI:** 10.64898/2026.08.31.26361844

**Authors:** Hend Jarras, Wilfried W. Bazié, Isalie Blais, Benjamin Goyer, Julien Boucher, Arielle Pakenham, Kimberly Dancause-Caron, Henintsoa Rabezanahary, Mathieu Thériault, Kim Santerre, Marc-André Langlois, Philippe A. Tessier, Jean-François Masson, Joelle N. Pelletier, Nicholas Brousseau, Denis Boudreau, Sylvie Trottier, Mariana Baz, Caroline Gilbert

## Abstract

Substantial inter-individual variation in susceptibility to viral infection persists despite widespread vaccination, and its immunological basis remains poorly understood. We investigated how innate and adaptive immune responses contribute to susceptibility to SARS-CoV-2 infection during the early COVID-19 pandemic. We compared two groups of vaccinated individuals who either remained uninfected or became infected during the first Omicron wave. Blood samples were collected at baseline and 24 weeks later. Peripheral blood mononuclear cells (PBMCs) and polymorphonuclear neutrophils (PMNs) were isolated and stimulated with the TLR7/8 agonist R848 to assess innate responses. PBMCs were stimulated with SARS-CoV-2 peptide pools and highly purified inactivated viruses (ancestral and Omicron BA.1) to evaluate adaptive immunity. Prior to infection, individuals in the infected group exhibited reduced CD4⁺ and CD8⁺ T cells proliferative responses, alongside with increased TNF production across all stimulation conditions, despite largely comparable immune phenotypes, indicating a pre-existing functional immune deficit. Following infection, T-cell proliferation and IFN-γ production were partially restored in response to viral antigens, although responses to Omicron BA.1 remained suboptimal. This functional deficit was accompanied by heightened inflammatory activity, including increased TNF and IFN-γ production, elevated anti-nucleocapsid IgG3 levels, higher frequencies of B cells and myeloid cells, reduced circulating interferon-inducible T-cell Alpha Chemoattractant (I-TAC) concentrations, and a modest impairment in PMN IL-8 responses. Notably, these alterations were detectable prior to infection and persisted thereafter, indicating that they represent determinants rather than consequences of viral infection. Importantly, beyond differences in the magnitude of immune responses, protection was associated with the degree of functional coordination within the humoral compartment, as reflected by the relationship between Spike-binding antibodies and neutralizing activity. Together, these results demonstrate that susceptibility to Omicron infection is linked to a pre-existing and persistent functional immune imbalance affecting both innate and adaptive arms of immunity.

**Highlights:**

- Pre-existing impairment in CD4⁺ and CD8⁺ T cell proliferative capacity identifies individuals at higher risk of Omicron infection.
- Susceptibility to Omicron infection is associated with a combined immune signature including altered cytokines (I-TAC, TNF), anti-N IgG3 antibodies, and cellular phenotypes.
- Omicron infected individuals displayed higher frequency of B cells and myeloid cells.
- Omicron infection partially restores T-cell function but fails to normalize responses to homologous viral stimulation.
- Extended 7-day proliferation assays using highly purified viral antigens enhance detection of functional immune deficits undetected by usual analysis alone such as ELISpot IFN-γ AIM assay or phenotype analysis.

## INTRODUCTION

The emergence of antigenically divergent SARS-CoV-2 variants has exposed important gaps in our understanding of the determinants of susceptibility to infection. Although vaccination and prior infection generate durable immune memory and substantially reduce the risk of severe disease, breakthrough infections remain common, particularly since the emergence of the Omicron lineage [1, 2]. These observations suggest that the presence of pre-existing immunity alone is insufficient to generate protection, and raise fundamental questions regarding the immune mechanisms that distinguish individuals who remain protected from those who become infected despite comparable vaccination histories.

The SARS-CoV-2 pandemic raised questions about viral evolution and the ability of the immune response to prevent severe disease and to protect against subsequent exposure [3–8]. Since the beginning of the pandemic, successive waves driven by distinct SARS-CoV-2 variants have occurred, each associated with varying levels of associated morbidity and mortality [9–13]. Some variants are less virulent, while others escape neutralization by antibodies induced by infections or immunizations [3–5, 7, 13]. The initial vaccine has decreased effectiveness against variants including B.1.351 (Beta), B.1.1.28 (Gamma), B.1.617.2 (Delta), and B.1.1.529 (Omicron) [9, 14–17]. The emergence of the Omicron variant at the end of 2021 marked a new chapter in the COVID-19 pandemic, introducing a less severe but more contagious virus that rapidly became the dominant variant worldwide [18–22]. Moreover, the acquired mutations in the spike protein challenged the previously developed infection- and vaccination-induced antibody responses [23–26]. Despite this inefficiency in antibody neutralization, immunity was not completely lost, as T cell responses seemed preserved, highlighting their importance for protection [27–30]. Omicron has also been shown to differentially induce the innate immune response compared to previous variants and to the ancestral strain, partly explaining its different pathogenicity [31–33]. Thus, it is important to understand the function of different components of the immune system and their roles in protection against this virus.

Much of the current understanding of SARS-CoV-2 immunity has focused on humoral responses. Neutralizing antibodies are used as correlates of protection against symptomatic COVID-19 and strongly predict vaccine efficacy at the population level [34, 35]. However, the extensive antigenic evolution of Omicron demonstrated that antibody-mediated protection is incomplete and highly susceptible to viral escape [2, 24]. Moreover, individuals with similar antibody levels frequently display markedly different susceptibility to infection, suggesting that additional qualitative and functional immune mechanisms contribute to protection.

Effective protection against viral infection emerges from the coordinated integration of innate and adaptive immune responses. Defining the mechanisms that sustain this immune coordination and understanding why it fails in certain individuals remain major challenges in human immunology. Innate and cellular immune responses are increasingly recognized as critical determinants of antiviral defense. Early sensing of viral RNA through pattern-recognition receptors, including TLR7 and TLR8, triggers interferon-dependent programs that limit viral replication and coordinate adaptive immunity [36–38]. Similarly, SARS-CoV-2-specific CD4^+^ and CD8^+^ T cells contribute to viral control and remain largely cross-reactive against Omicron variants despite extensive spike mutations [39, 40]. Yet, most studies have examined these immune compartments separately, and the extent to which their functional integration influences susceptibility to infection remains poorly understood.

As first-line responders, neutrophils attempt to eliminate pathogens through the production and release of reactive oxygen species and granule contents rich in proteases, metalloproteins, membrane receptors and myeloperoxidase, or by producing inflammatory cytokines such as IL-6, TNF, and IL-8 [41]. IL-8 in particular plays a pivotal role in recruiting immune cells, including neutrophils and lymphocytes, especially during viral infections [42, 43]. Polymorphonuclear neutrophils (PMN) and mononuclear myeloid cells (dendritic cells and monocytes) detect viruses by means of pattern recognition receptors (PRRs) and initiate inflammatory and immune responses [44–46]. Toll-like receptors (TLR) 7/8 recognize guanosine- and uridine-rich (GU-rich) sequences of single-stranded RNA (ssRNA) viruses. In the case of SARS-CoV-2, those sequences are named SARS-CoV-2-associated molecular patterns (SAMPs) [47, 48]. A previous study on SARS-CoV has shown that this GU-rich motif may significantly contribute to the cytokine storm and to the dysregulation of the innate immune response [49]. TLR7/8 activation orchestrates specific immune responses through several pathways [50, 51]. For example, IFN response during SARS-CoV-2 infection may vary substantially between patients and may differ according to disease severity. An early IFN response by innate immune cells can be protective against infection and severe disease [12, 41, 52, 53]. These IFN responses appear to play a role, first by limiting viral entry, translation, replication and assembly, and second, by coordinating and accelerating the development of adaptive immunity [12, 41, 52, 53]. However, coronaviruses, including SARS-CoV-2, have developed several evasion strategies to counteract this host defense including limiting type I and type III IFN production at post-transcriptional levels, and altering proinflammatory cytokine expression [54–56].

Reinfection, or evolution to severe forms of COVID-19, may be due to the failure these innate immunity mechanisms and/or to the development of an aberrant acquired immune response [12, 57, 58]. Exposure to the virus, or immunization, elicits adaptive immunity and leads to the establishment of immunologic memory [17, 59]. SARS-CoV-2 infection induces polyclonal humoral and cellular responses targeting multiple viral proteins [60, 61]. In COVID-19 patients, longitudinal analysis shows durable and broad immune memory after infection, with persisting neutralizing antibodies and memory B and T cells that recognize distinct viral epitope regions [62]. The virus-specific T-cell responses are reported to increase with disease severity, suggesting a deficiency in adaptive immunity [63, 64]. Likewise, vaccination also induces rapid antigen-specific CD4^+^ T-cell responses in naive subjects, whereas CD8+ T-cell responses develop gradually and are variable in magnitude [65]. The CD4^+^ (Th1 and Tfh) T-cell responses are efficiently generated following primary vaccination, and are strongly correlated with post-boost neutralizing antibody levels and CD8^+^ T-cell responses [65].

Neutralizing antibodies in particular are of great importance as they act by blocking viral entry into cells [66]. A recent study on SARS-CoV-2 have shown that both IgG1 and IgG3 dominate the humoral response during infection, with IgG3 often appearing early and correlating with strong effector functions, whereas IgG1 is more prevalent after vaccination and is associated with sustained neutralization capacity against variants, including Omicron [67]. Together, these subclasses orchestrate a synergistic response that combines direct neutralization with immune- mediated clearance, and is therefore relevant in relation to protection against infection.

Characterizing the factors associated with infection and protection despite previous exposure to SARS-CoV-2 (through infection or vaccination) is relevant at the individual, clinical, epidemiological, and public health levels, as it may help identify vulnerable populations, improve risk stratification, optimize vaccination strategies, and strengthen preparedness for future outbreaks. For this purpose, more comprehensive quantitative and qualitative analyses of both innate and adaptive immunity are needed to understand how some individuals are more susceptible to infection.

Here, we performed longitudinal multidimensional immune profiling of vaccinated individuals exposed during the first Omicron BA.1 wave. By integrating humoral, innate, cellular, and functional immune analyses before and after viral exposure, we sought to define immune states associated with susceptibility or resistance to breakthrough infection. Our findings reveal that susceptibility to Omicron infection is not explained by antibody levels alone but is associated with a pre-existing state of immune discoordination characterized by impaired T-cell fitness, altered interferon-associated signaling, defective neutrophil responsiveness, and distinct humoral immune features. Together, these data identify immune coordination across innate and adaptive compartments as a key determinant of protection against SARS-CoV-2 infection.

## RESULTS

### Clinical and Demographic Data

This work consisted in the study of the innate and adaptive immune response in individuals who developed or not SARS-CoV-2 infection during the period of observation. Blood samples were collected at baseline (V1) and 24 weeks later (V3). V1 took place before Omicron predominance, between June and October 2021, while V3 occurred during Omicron predominance from December 2021 to April 2022 (**Fig. 1**). The visits and vaccination timelines, as well as the number of SARS-CoV-2 infections per month are represented in **figure 1B**. All participants were initially free of infection and fully vaccinated (≥ 2 doses) by V3. Only one participant had received a third vaccine dose at V1. The non-infected group included 31 participants without evidence of SARS-CoV-2 infection at V3 while the infected group consisted of 44 individuals with evidence of SARS-CoV-2 infection at V3 (**Fig. S1).** Median age was 34.0 years (IQR 22.0-51.0) in the non-infected group and 33.0 years (IQR 25.3-47.8) in the infected group (p=0.5980; **Table 1**). Females represented 58.1% of the non-infected individuals and 68.1% of infected individuals (p=0.4651). Moreover, according to an assessment of the level of exposure to the SARS-CoV-2 virus at V1 (**Table S1**), non-infected and infected individuals had a low to medium level of exposure, with no significant difference between both groups (p=0.2382). Most subjects receiving an mRNA vaccine as their first (81.4%) and second dose (96.7%). At V3, infected participants had a significantly longer elapsed time between the last vaccine dose received and their visit compared to V1 (**Fig. S2**). Classification according to their workplace (bar or restaurant, grocery store or hardware store), level of education, BMI, comorbidities and smoking/vaping are also presented in **Table S2**. The participants’ home and workplace zip codes were also compared and did not show a concentration in a specific region for both groups (**Fig. S3**). Together, these characteristics indicate that the two groups were comparable in terms of general demographic and clinical characteristics.

**Figure 1.**
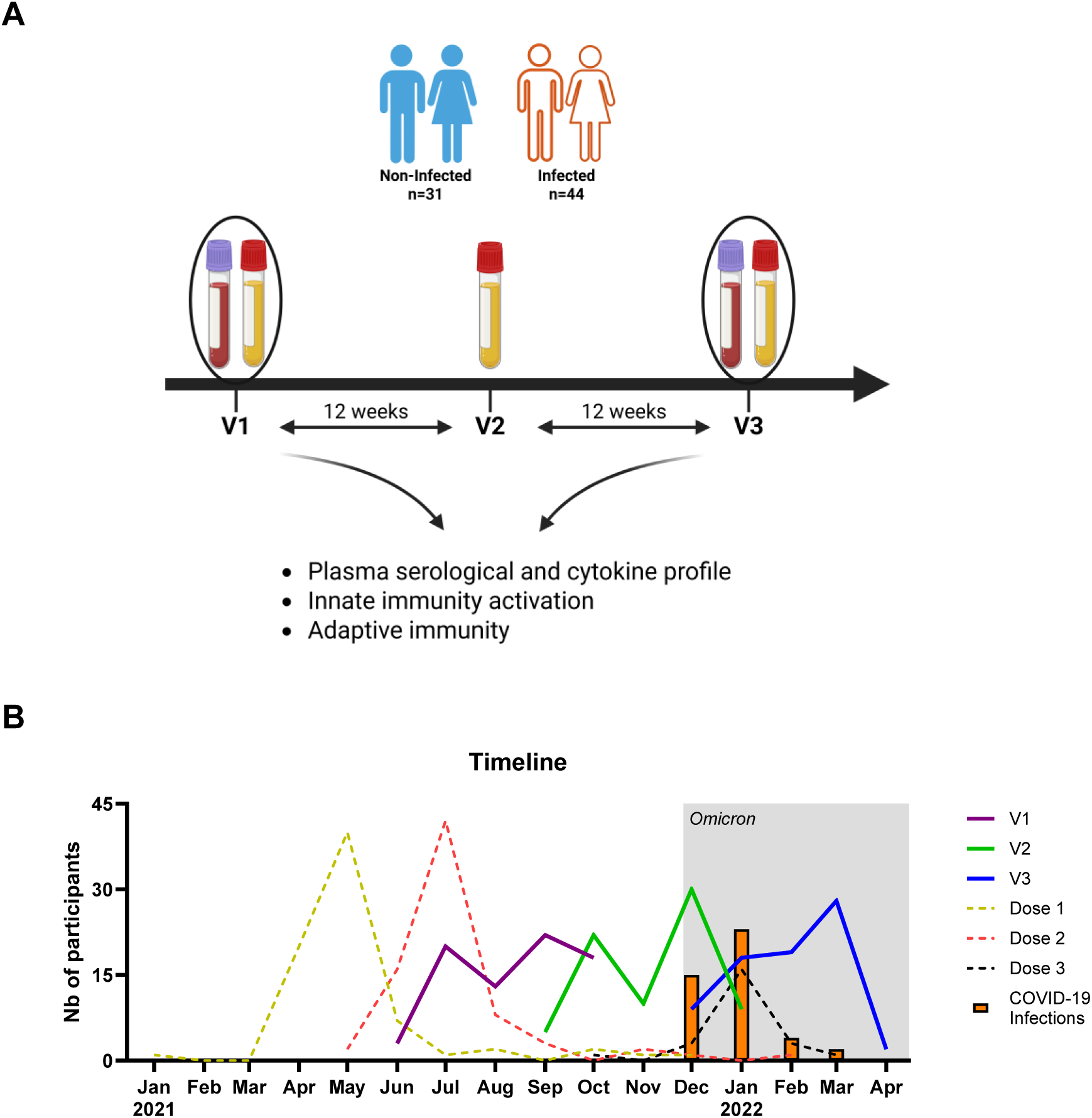
Schematic study description. **A)** Brief graphical representation of the study. Description of visit 1 (V1) and visit 3 (V3) at 24 ± 2 weeks interval. Blood was collected before (V1) and during (V3) Omicron predominance. Serum, plasma, PBMCs, and PMNs were isolated for subsequent analyses. **B)** Number of participants at each visit that took place from June 2021 to April 2022 (full lines), as well as the vaccine doses received starting January 2021 (dotted lines). The shaded area represents SARS-CoV-2 Omicron variant predominance. The number of SARS-CoV-2 infections per month are represented by orange colored bars. (A) was created with Biorender. See figure S10.

**Table 1.**
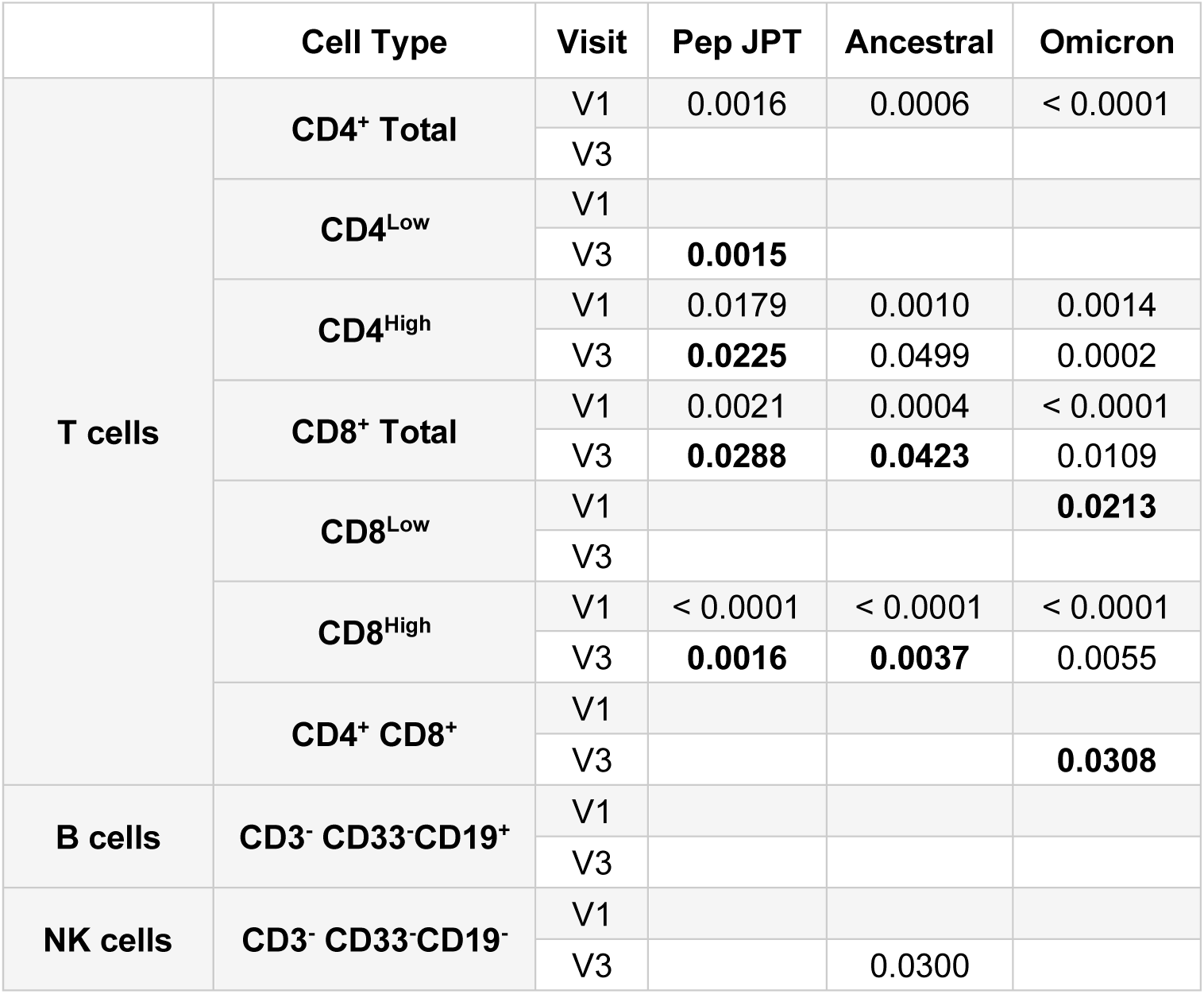
Proliferation assay reveals functional immune defect in infected participants. Comparison between the non-infected (n=18-28) and infected groups (n=27-38) at V1 and at V3. Values in bold represent an increase in the proliferation index of the infected group compared to the non-infected group, while values in red represent a decrease. Data were analized with Mann Whitney two-tailed unpaired t test.

### Vaccinal and Hybrid Immunity Evaluation in Non-Infected and Infected Individuals

Levels of anti-receptor binding domain (RBD), anti-spike (S), and anti-nucleocapsid (N) antibodies from the ancestral virus (**Fig. 2A**) were measured in individual sera as already described [68]. At V1, no significant difference was found in total IgG antibody levels between both groups (**Fig. 2B- D**), indicating that infected and non-infected participants had developed a similar vaccinal response. At V3, there was a significant increase in anti-S, anti-RBD and anti-N antibodies in infected individuals, demonstrating hybrid immunity [69–71]. These results were confirmed using an in-house ELISA (**Fig. S4**). While no differences were detected in anti-RBD and anti-S IgG3s, anti-N IgG3 antibody levels were moderately higher at V1 and significantly increased at V3 in infected participants (p<0,001, **Fig. 2E-G**). These data confirmed infection status of participants at V3 and corroborate their hybrid immunity status.

**Figure 2.**
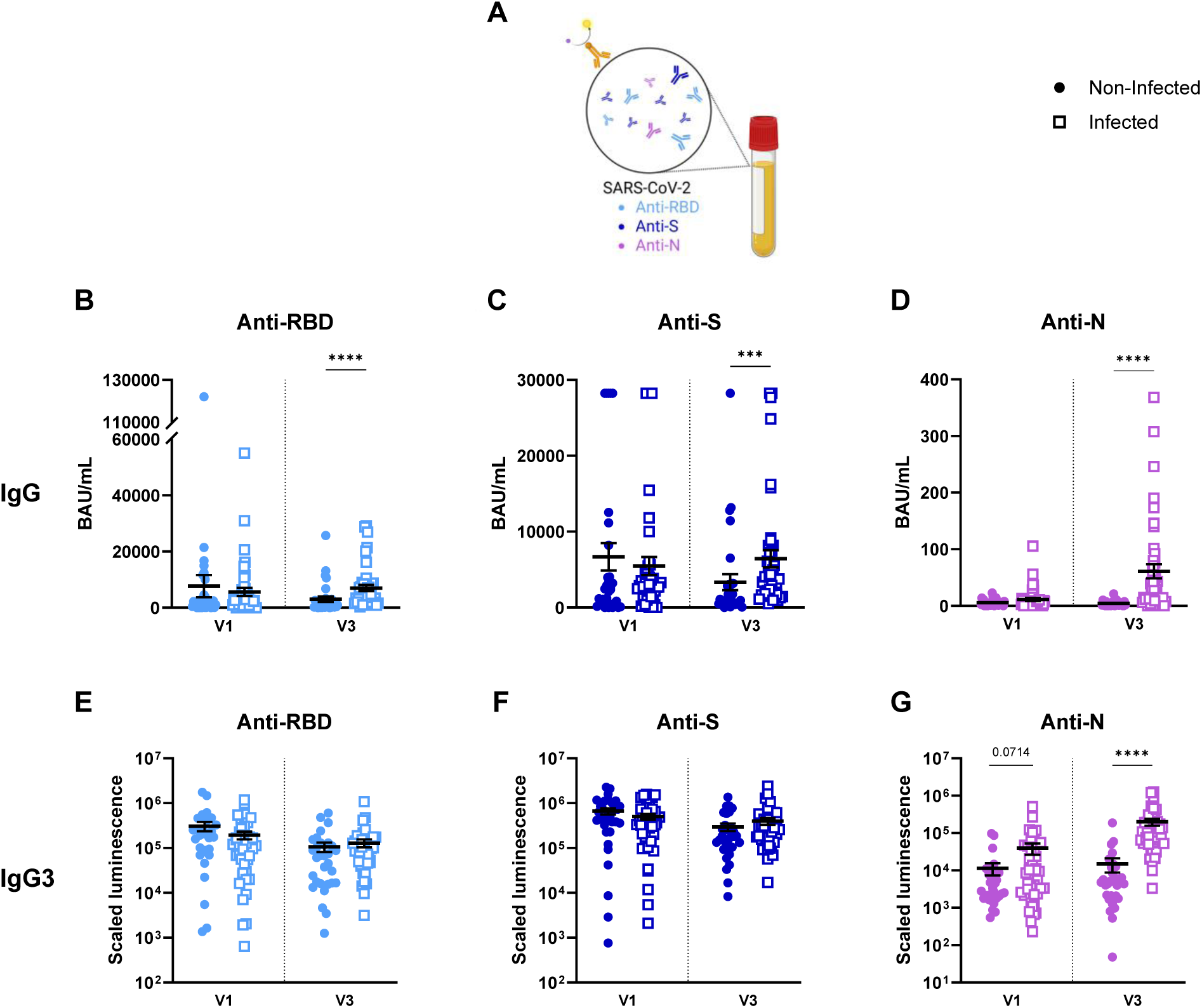
Serum antibody levels confirm infection at V3. Serum antibody levels in non-infected (full dots; n=31) and infected (empty squares; n=44) subjects during their first (V1) and third (V3) visits. **A)** Schematic representation of the plasma antibodies measured through chemiluminescence. Total IgG antibodies against **B)** the receptor binding domain (RBD) (light blue), **C)** the spike (S) protein (dark blue), and **D)** the nucleocapsid (N) protein (purple). Relative IgG3 concentrations of **E)** anti-RBD, **F)** anti-S, and **G)** anti-N antibodies. Each dot represents a participant. Data are shown as mean ± SEM and were analized with Mann-Whitney two-tailed unpaired t test. *** p < 0.001; **** p < 0.0001. BAU: binding antibody units. (A) was created with Biorender. See figures S3 & S4.

The neutralization capacity of serum to inhibit viral replication of different variants was characterized to assess antibody functionality (**Fig. 3A**) [72, 73]. At V1 (before the predominance of Omicron BA.1) infected and non-infected participants had similar levels of neutralizing antibodies against the ancestral, Delta and Omicron BA.1 strains (**Fig. 3B-D**). Notably, while the majority of participants in both groups developed neutralizing antibodies against the ancestral and the Delta strains, only 12.9% of non-infected and 9.1% of infected participants had neutralizing antibodies against the Omicron BA.1 variant at V1. At V3, serums from infected individuals showed improved neutralizing capacity against the three strains compared to the non-infected group. Collectively, these data show that the neutralization capacity of non-infected and infected participant sera are clearly distinct and highlight the importance of hybrid immunity in protecting against SARS-CoV-2.

**Figure 3.**
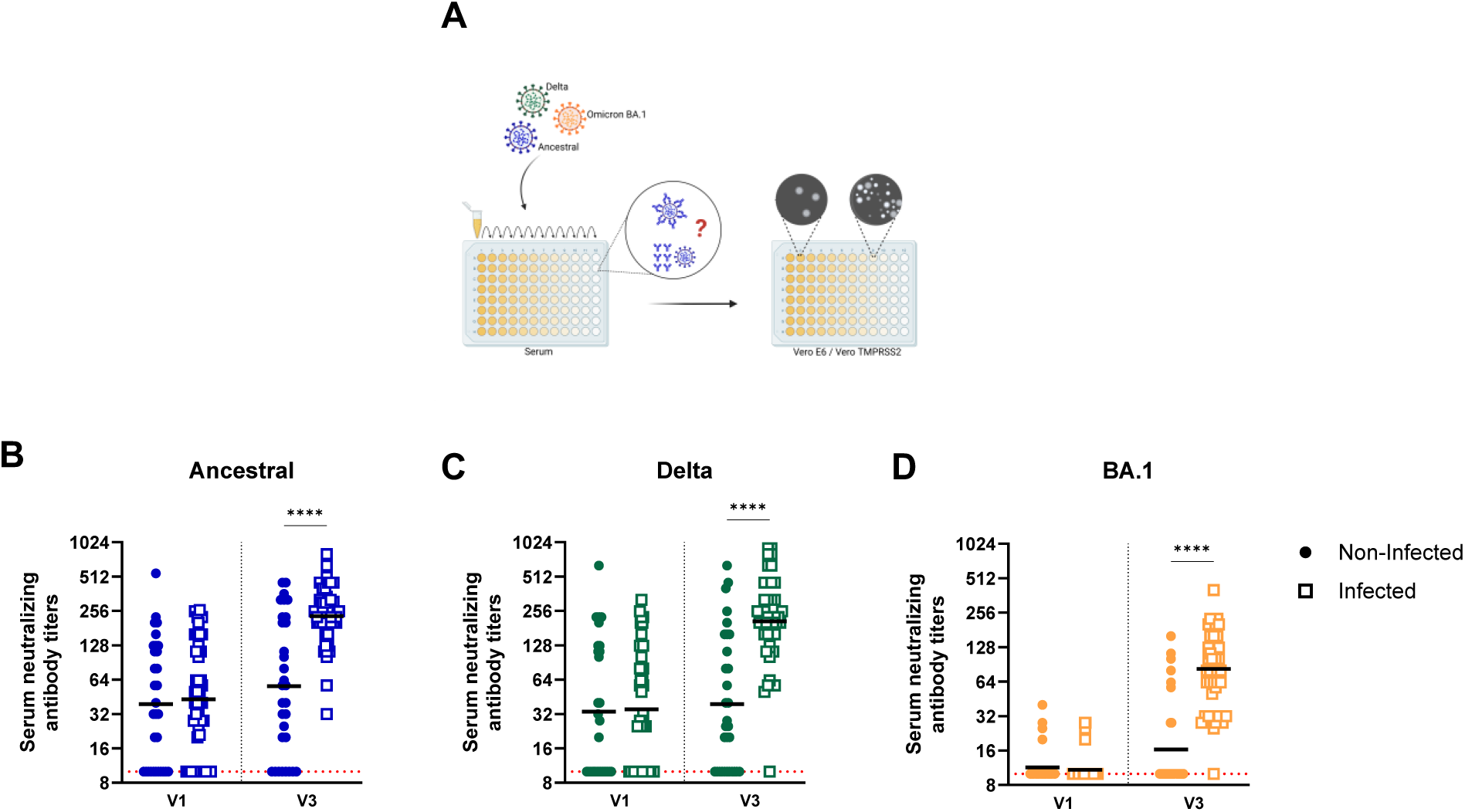
Hybrid immunity confers better serum neutralization capacity. Microneutralization assay in non-infected (n=31) and infected (n=44) individuals before (V1) and during (v3) Omicron predominance. Each dot represents a single participant. **A)** Schematic representation of the microneutralization test. Neutralization capacity of serum against **B)** ancestral virus. **C)** Delta variant and **D)** Omicron BA.1 variant. Each dot represents a participant. Data are shown as geometric mean and were analized with Mann-Whitney two-tailed unpaired t test. **** p < 0.0001. (A) was created with Biorender.

Since antibody-mediated protection depends not only on the magnitude of antigen-specific responses but also on their functional capacity, the relationship between spike-binding antibodies and viral neutralization was analysed (**Fig. S5**) to provide additional insight into immune mechanisms associated with susceptibility or resistance to Omicron BA.1 infection. To address this question, we examined correlations between neutralizing activity against ancestral and anti-S or anti-RBD IgG and IgG3 antibody levels at baseline (V1), as well as following the Omicron BA.1 wave (V3) in participants who became infected and those who remained uninfected (**Fig. S5)**. Interestingly, the relationship between binding and functional antibody responses differed markedly according to infection outcome. At baseline (V1), neutralizing activity correlated significantly with both anti-S (p=0.0473) and anti-RBD (p=0.0207) IgG antibody levels in participants who subsequently became infected, indicating that neutralization was largely driven by spike-specific humoral responses (**Fig. S5A, C**). Following Omicron BA.1 infection (V3), these correlations were lost, suggesting a decoupling between antibody binding magnitude and neutralizing function (**Fig. S5B, D**). In contrast, significant correlations emerged in individuals who remained uninfected (p<0.0001), revealing a tighter association between spike-specific antibody levels and neutralization capacity. As for IgG3, significant correlations were seen for both groups at V1 for anti-S (non-infected p=0.0006, infected p=0.0008) and anti-RBD (non-infected p=0.0004, infected p=0.0009) antibodies (**Fig. S5E, G**), but the group that was infected lost that correlation at V3 (**Fig. S5F, H**). These findings suggest that Omicron infection qualitatively reshapes the humoral immune response and that protection from infection may be associated with a more coordinated relationship between antibody binding and antiviral function.

### Pre-Infection Cytokine Profiling and Alterations Following Omicron Infection

We next compared the mean cytokine levels in platelet-free plasma of each subgroup to the whole cohort [74]. Most cytokines did not show significant differences between groups at V1 (**Fig. 4**). However, concentrations of I-TAC/CXCL11, a cytokine involved in interferon-mediated immune responses [75], were significantly lower at baseline in individuals who later became infected compared to the overall cohort. In contrast, comparison of non-infected and infected participants at V3 revealed significant changes in multiple cytokines between each group. Notably, a greater number of cytokines were downregulated at V3 in the infected group (**Fig. S6**), suggesting that Omicron infection induced a marked dysregulation of cytokine production. This contrasts with calprotectin which was increased at V3 in non-infected and infected groups. These results show that the concentration of cytokines and inflammatory markers in plasma were decreased by infection.

**Figure 4.**
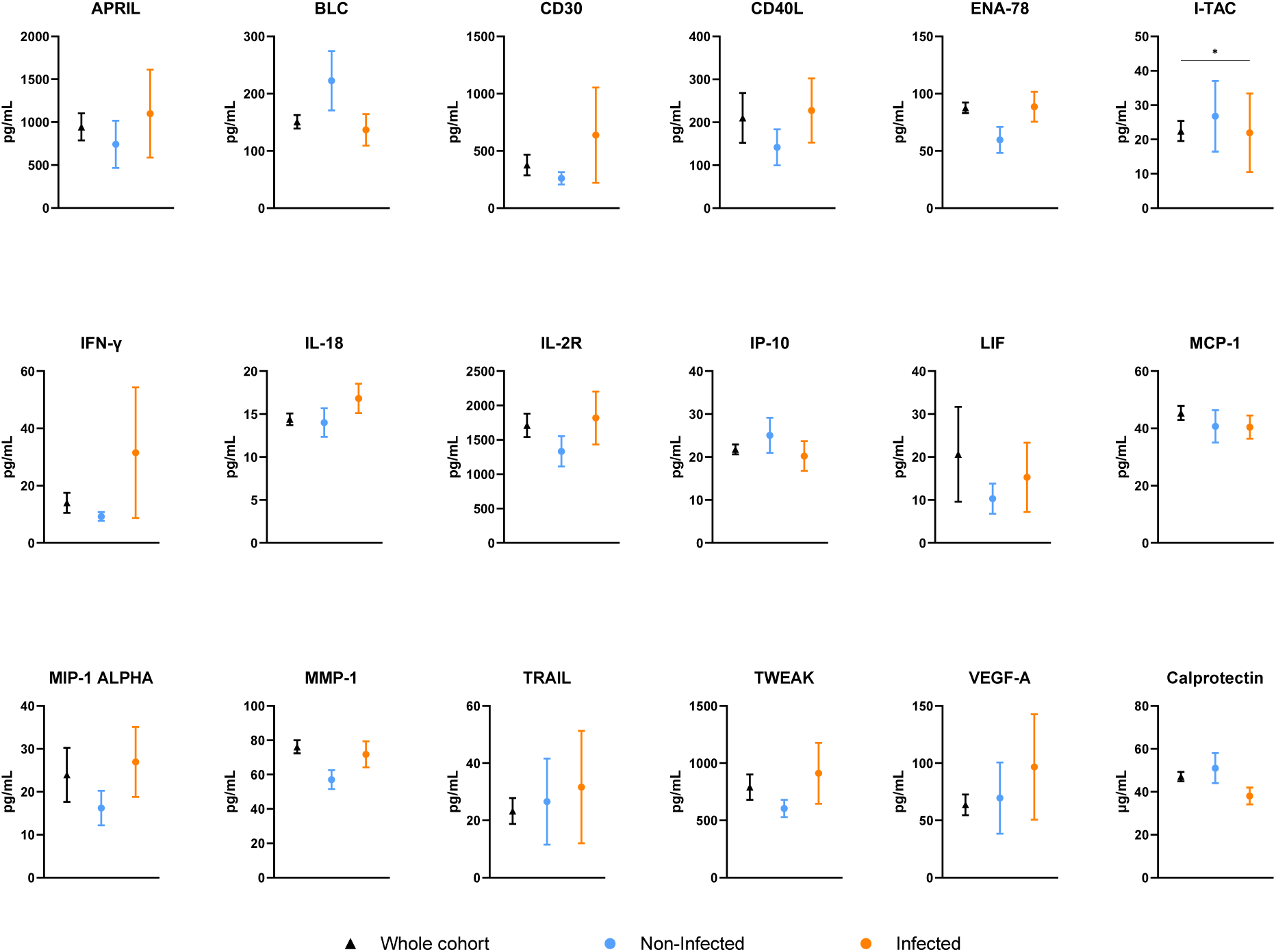
Reduced I-TAC levels in infected participants at V1. Mean cytokine levels before (V1) Omicron predominance, for the non-infected (n=31) and infected (n=44) groups compared to the whole cohort (n=304) mean. Data are shown as mean ± SEM and were analyzed with Kruskal-Wallis test with Dunn’s multiple comparisons test. * p < 0.05. See figure S5.

### Innate Response Pre- and Post-Infection

As PMNs and PBMCs are important producers of cytokines and inflammatory molecules, these cells have been stimulated with R848, a TLR7/8 agonist, to assess the innate response of participants (**Fig. 5A, 6A**). Cytokine levels (IL-8, TNF and IFN-γ) were then measured in the supernatant by ELISA. As shown in **Figure 5B-C**, although the infected group consistently exhibited slightly lower IL-8 levels secreted by PMNs at V1, a significant difference in released IL- 8 between the non-infected and infected groups was observed at V3, but only following stimulation with 1 µg/mL R848. Similarly, IL-8 levels were slightly lower in supernatants of PBMCs from infected participants, but no significant difference was observed for both visits (**Fig. 6B-C**). Further, when comparing IL-8 production between V1 and V3 in each group, no significant difference was observed in the supernatants of R848-stimulated PMNs (**Fig. S7**), or PBMCs of non-infected individuals (**Fig. S7A**). However, IL-8 production in the infected group at V3 (post-infection) was significantly higher than prior infection (**Fig. S8B**).

**Figure 5.**
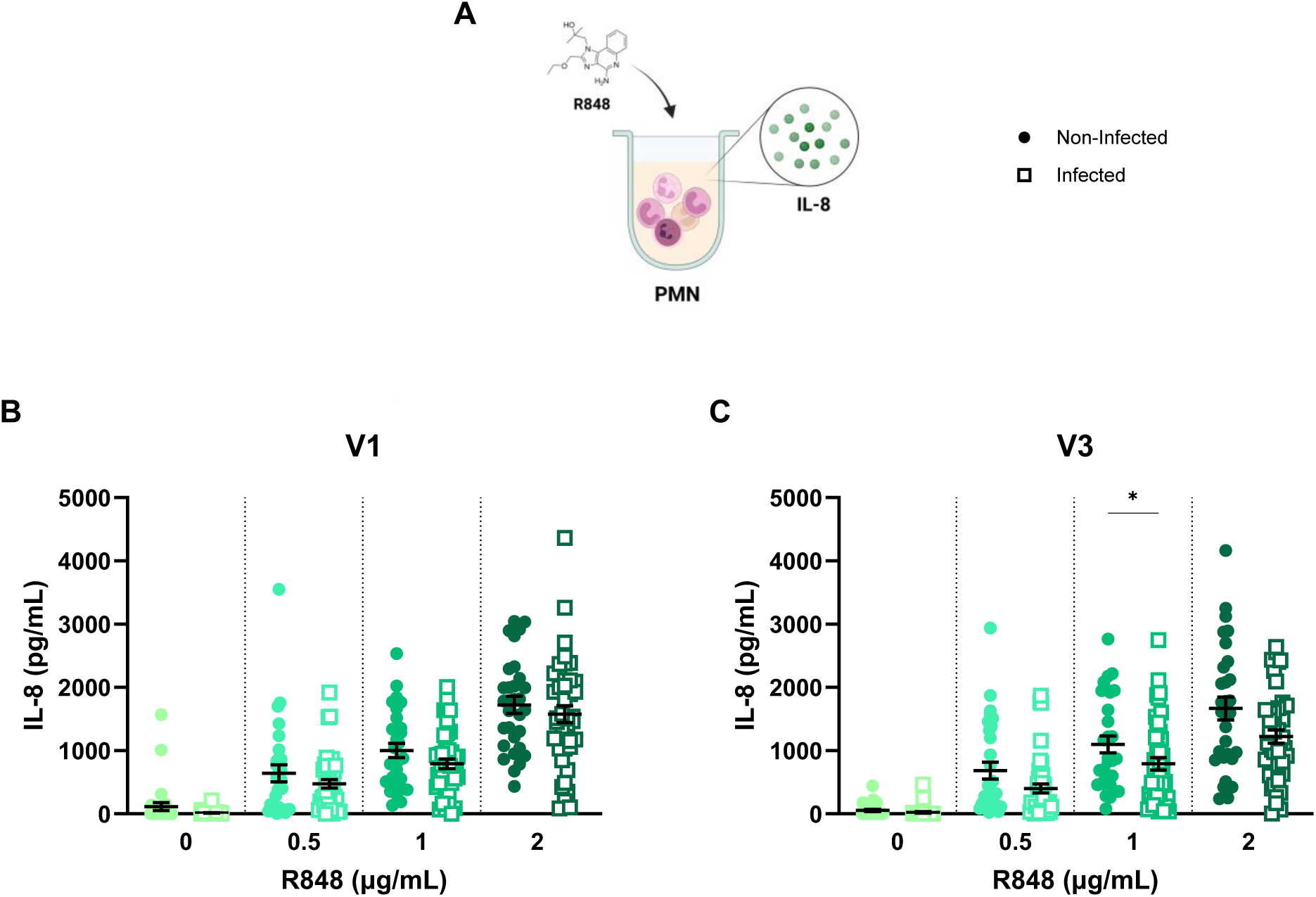
PMN immune response is weaker in infected individuals. Measures of IL-8 levels produced by PMNs in non-infected (full dots) and infected (empty squares) groups after 24h of stimulation with different concentrations of R848. **A)** PMNs were incubated 24h with R848, producing and releasing IL-8 in the supernatant. **B)** IL-8 responses of non-infected (n=30) and infected (n=42) participants at V1. **C)** IL-8 responses of non-infected (n=30) and infected (n=44) participants at V3. Each dot represents a participant. Data are shown as mean ± SEM and were analized with Mann-Whitney two-tailed unpaired t test. * p < 0.05. (A) was created with Biorender. See figure S6.

**Figure 6.**
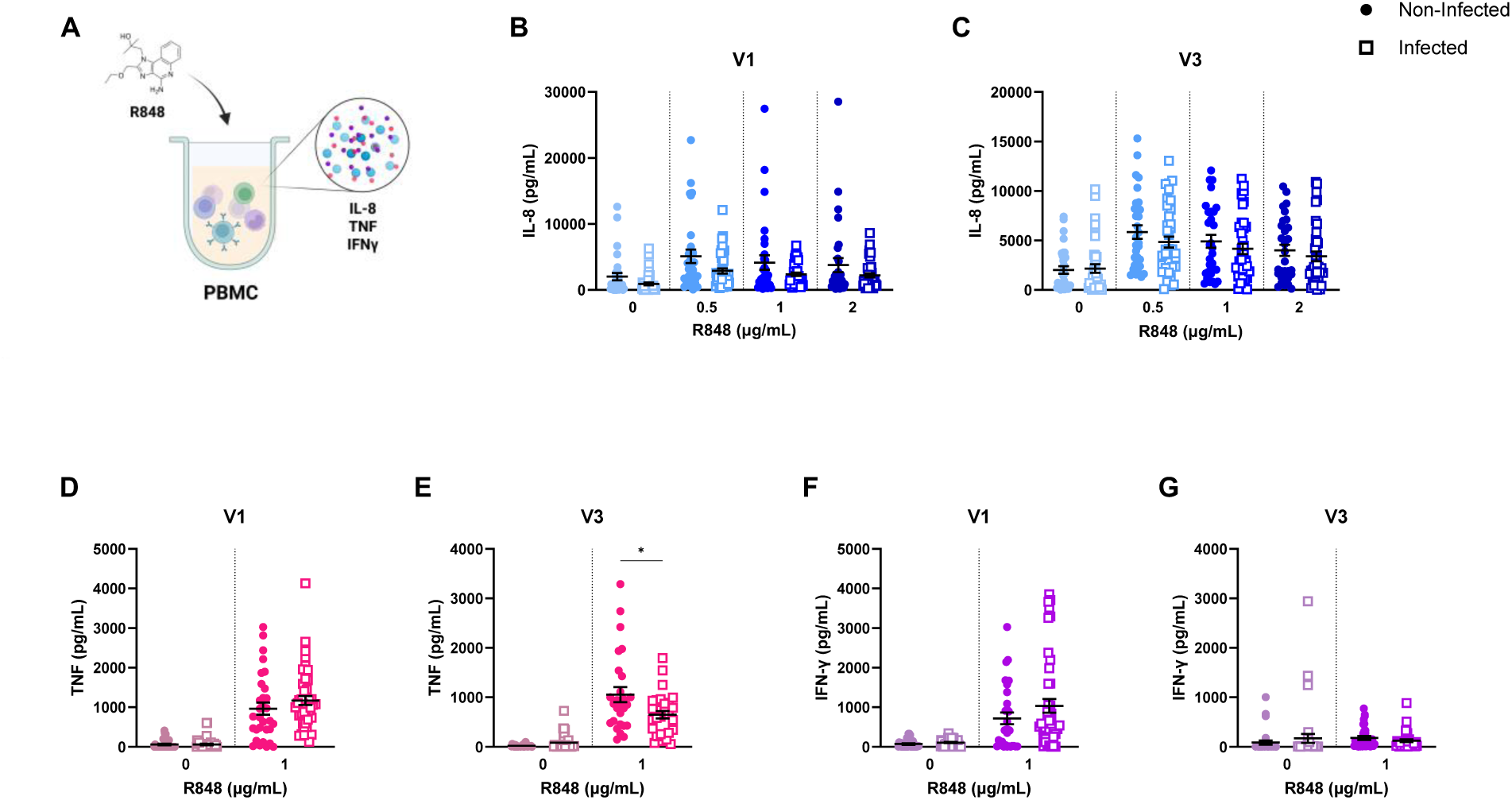
PBMC cytokine profile is slightly modified following Omicron BA.1 infection. Measures of cytokine levels produced by PBMCs in non-infected (full dots) and infected (empty squares) groups after 24h of stimulation with different concentrations of R848. **A)** PBMCs were incubated 24h with R848, producing and releasing IL-8, TNF and IFN-γ in the supernatant. **B)** IL- 8 responses of non-infected (n=31) and infected (n=43) participants at V1. **C)** IL-8 responses of non-infected (n=31) and infected (n=39) participants at V3. **D)** TNF responses of non-infected (n=31) and infected (n=44) participants at V1. **E)** TNF responses of non-infected (n=27) and infected (n=32) participants at V3. **F)** IFN-γ responses of non-infected (n=31) and infected (n=44) participants at V1. **G)** IFN-γ responses of non-infected (n=31) and infected (n=38) participants at V3. Each dot represents a participant. Data are shown as mean ± SEM and were analized with Mann-Whitney two-tailed unpaired t test. * p < 0.05. (A) was created with Biorender. See figures S7 & S8.

PBMCs from the infected group produced significantly less TNF in response to R848 at V3 than PBMCs from the non-infected group (**Fig. 6D-E**). In contrast, IFN-γ was produced at similar levels by PBMCs from both groups (**Fig. 6F-G**). While no difference was observed between visits in non- infected participants, a decrease in TNF levels at V3, compared to V1, was observed in supernatants from PBMCs of infected participants (**Fig. S9A-B**). In addition, the non-infected group showed a decrease in IFN-γ’s secretion at V3 following stimulation with 1 µg/mL R848 (**Fig. S9C**), while supernatants from PBMCs of the infected group showed significantly increased levels at V3 at rest and decreased levels following stimulation (**Fig. S9D**). These findings indicate that prior to infection, the innate immune response exhibits a comparable profile across the studied groups, and that Omicron infection induces a long-lasting modification of the cytokine profile of both PMNs and PBMCs.

### Multiple Components of Specific Immune Response Pre- and Post-Infection

Next, lymphocyte proliferation was examined in participants’ PBMCs collected before (V1) and during Omicron predominance (V3) to investigate the cellular immune response. Immune cell phenotypes were first determined before stimulation (**Fig. S10**) and compared between non-infected and infected groups. A larger proportion of B cells (**Fig. S10F**) was observed in blood from infected participants at V1 and V3 (p<0.001), which highlights increased antibody-producing cells. For all other cell types, the proportion of live cells per individual was similar in infected and non-infected groups before stimulation. After stimulation, the percentage of myeloid cells, compared to all live cells at V1 and V3, was higher in infected individuals (**Fig. 7**). The percentages of T cells, and more specifically CD4^+^ T cells, were relatively lower in infected participants at V1 and V3 in the absence of stimulation, and after seven days of stimulation with both inactivated viruses (**Fig. 8**). There was a higher proportion of B cells in blood from the infected group in all conditions for both visits. Other cell phenotypes were comparable between the two groups (**Fig. 8**). These data reveal a marked immune imbalance associated with Omicron susceptibility, characterized by a reduced CD4⁺ T cell response and concurrent expansion of myeloid and B cell compartments, suggesting an altered cellular landscape that may compromise antiviral defense.

**Figure 7.**
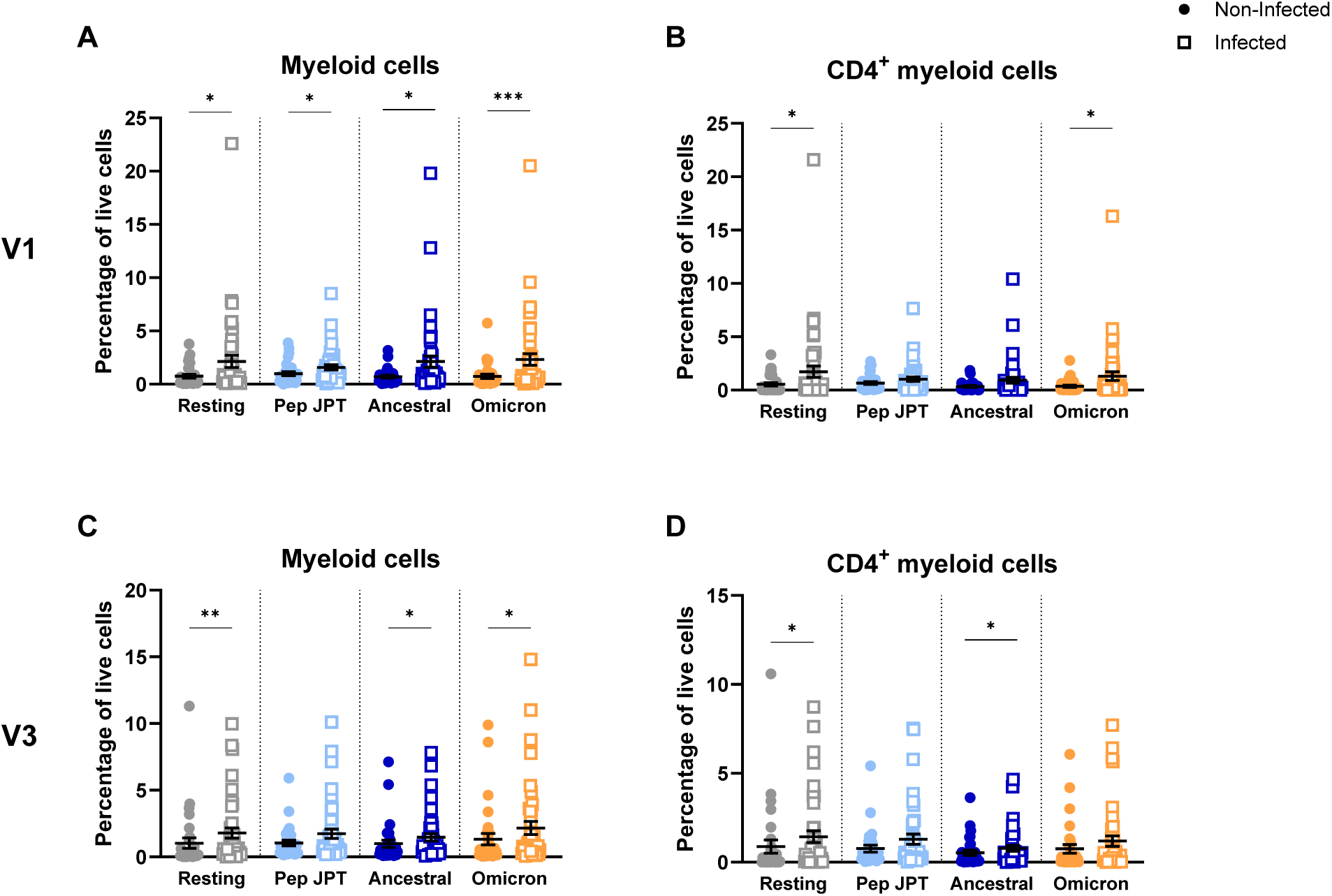
Infected individuals retain a higher proportion of myeloid cells following 7-day assay both pre- and post-infection. Proportion of live cells at day 7 of the proliferation assay before (V1) and during (V3) Omicron predominance. **A)** Myeloid cells (CD33^+^) and **B)** CD4^+^ myeloid cells (CD33^+^ CD4^+^) in non-infected (n=30) and infected (n=44) participants at V1, and **C)** myeloid cells and **D)** CD4^+^ myeloid cells in non-infected (n=31) and infected (n=42) individuals at V3. Each dot represents a participant. Data are shown as mean ± SEM and were analized with Mann-Whitney two-tailed unpaired t test. * p < 0.05; ** p < 0.01; *** p < 0.001. Resting: no stimulation; Pep JPT: pool of peptides derived from the SARS-CoV-2 Spike glycoprotein; Ancestral: inactivated ancestral strain of SARS-CoV-2; Omicron: inactivated sublineage BA.1 Omicron variant of SARS-CoV-2. See figure S9.

**Figure 8.**
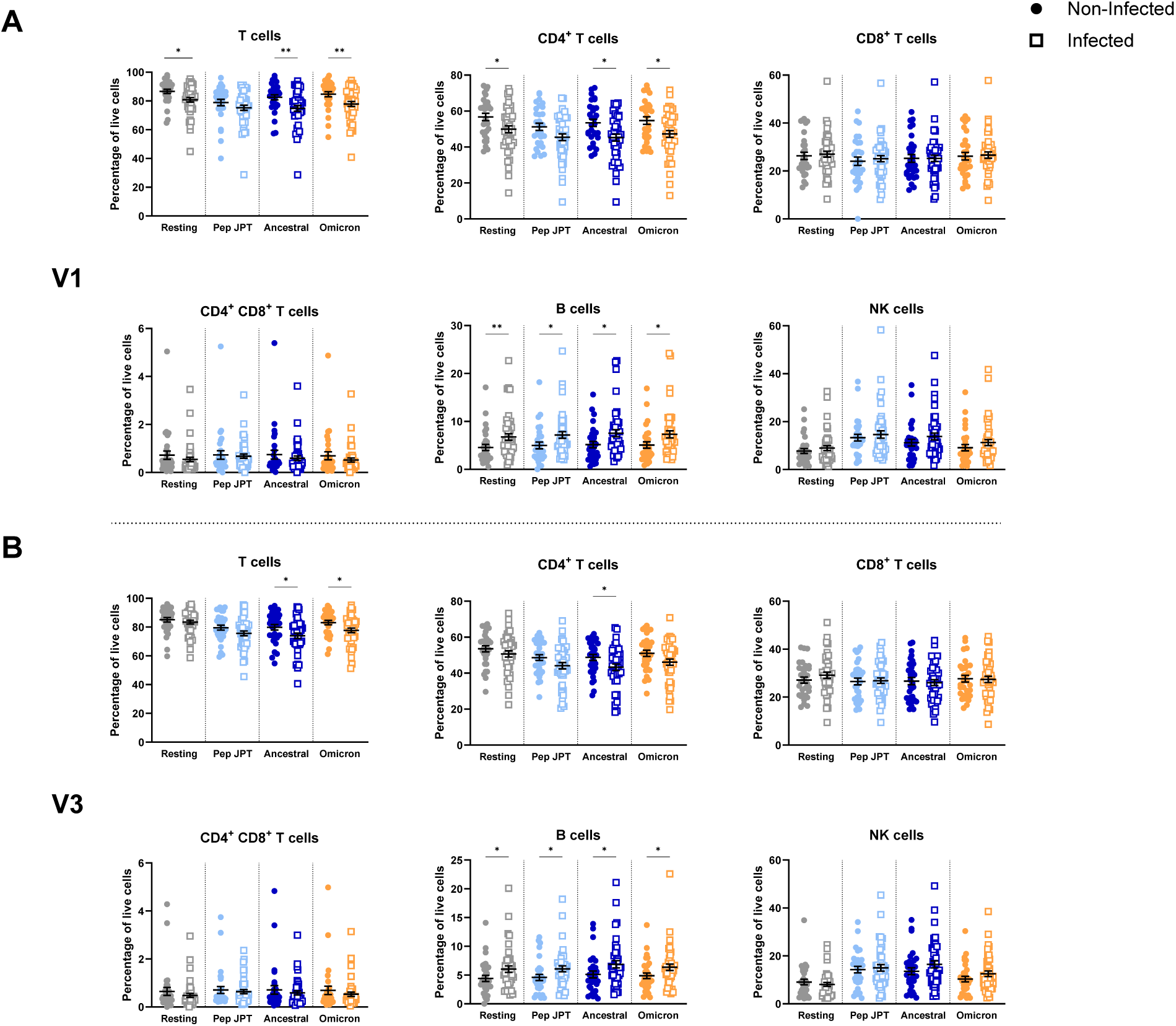
Infected individuals have lower CD4^+^ T cell and higher B cell proportion compared to non-infected. Proportion of live cells at day 7 of the proliferation assay for each of the following phenotypes: T cells (CD33^-^ CD3^+^), CD4^+^ T cells (CD33^-^ CD3^+^ CD4^+^), CD8^+^ T cells (CD33^-^ CD3^+^ CD8^+^), double positive T cells (CD33^-^ CD3^+^ CD4^+^ CD8^+^), B cells (CD33^-^ CD3^-^ CD19^+^), and NK cells (CD33^-^ CD3^-^ CD19^-^). **A)** V1 (non-infected n=30 and infected n=44), and **B)** V3 (non-infected n=31 and infected n=42). Each dot represents a single participant. Data are shown as mean ± SEM and were analized with Mann-Whitney two-tailed unpaired t test. * p < 0.05; ** p < 0.01. Resting: no stimulation; Pep JPT: pool of peptides derived from the SARS-CoV-2 Spike glycoprotein; Ancestral: inactivated ancestral strain of SARS-CoV-2; Omicron: inactivated sublineage BA.1 Omicron variant of SARS-CoV-2.

The proliferation index, measured by CFSE cell staining, represents the average number of divisions of the cells that specifically responded to the stimulation [76]. The significant p-values of the comparisons between the non-infected and infected groups at V1 and at V3 for each relevant cell type and condition of stimulation are presented in **Table 1**. **Table 2** shows the significant p-values of the comparisons between V1 and V3 for each study group separately. In the comparison between both groups (**Table 1**), we observed that cells from participants in the infected group had a lower (red) proliferation index than those from the non-infected group at V1, for all conditions of stimulation, for both CD4^+^ and CD8^+^ T cells. This suggests that subjects in the infected group had a weaker cellular response before infection. An opposite trend was observed at V3, with a stronger response (green) in the infected group following stimulation with the peptide mix and the inactivated ancestral virus in CD4^+^ and CD8^+^ T cells, suggesting that the infection improved the T cell response. Responses to the Omicron BA.1 virus remained lower for CD4^+^ and CD8^+^ T cells in infected participants compared to non-infected participants. However, CD4^+^ CD8^+^ (double positive) T cells, an activated phenotype of T cells [77], had a significantly higher proliferation index for infected participants at V3 when stimulated with the inactivated Omicron BA.1 virus. Triple negative cells, which we presume to be NK cells, had a significantly lower proliferation index in infected subjects at V3. We observed few changes in the non-infected group when comparing the proliferation index between V1 and V3 (**Table 2**). For the infected subjects, there was an increase in the proliferation index after infection mostly for CD4^+^ and CD8^+^ T cells under all conditions. The CD4^+^ CD8^+^ T cells were more proliferative in infected participants at V3 following stimulation with the inactivated Omicron BA.1 virus. There was also an increase in the proliferation index of B cells for both groups at V3 following stimulation with the inactivated ancestral virus. Lastly, the triple negative cells showed stronger proliferation at V3 in infected individuals. We also compared the expansion indices, which represent the fold expansion of the whole culture, in order to understand the overall response of cells [76]. Results showed that PBMCs (CD4^+^, CD8^+^ and NK cells) harvested from the infected group were less prone to divide, (**Table S3-4**). B cells were the only cell type to show no difference in expansion both between groups and visits. Interestingly, double positive CD4^+^ CD8^+^ T cells had a significantly higher expansion index in infected subjects at V3 when stimulated with the inactivated Omicron BA.1 virus. Data also show that CD4 ^High^ and CD8^High^ T cells demonstrated a decreased expansion index at V3 mainly in the infected group, suggesting a reduced capacity of the whole PBMC culture to be activated and proliferate after SARS-CoV-2 infection. These results reveal a functional immune defect preceding infection compared to participants who remained uninfected, characterized by an impaired T cell proliferative capacity despite preserved phenotypic distributions. This deficit is partially restored after infection but remains suboptimal, particularly in response to Omicron, highlighting a persistent functional gap in antiviral immunity. Importantly, the use of a 7-day proliferation assay combined with highly purified inactivated viral stimuli provides a robust and sensitive framework to capture these subtle yet biologically meaningful differences, underscoring its added value in identifying functional correlates of susceptibility that are not detectable by phenotypic analysis alone.

**Table 2.**
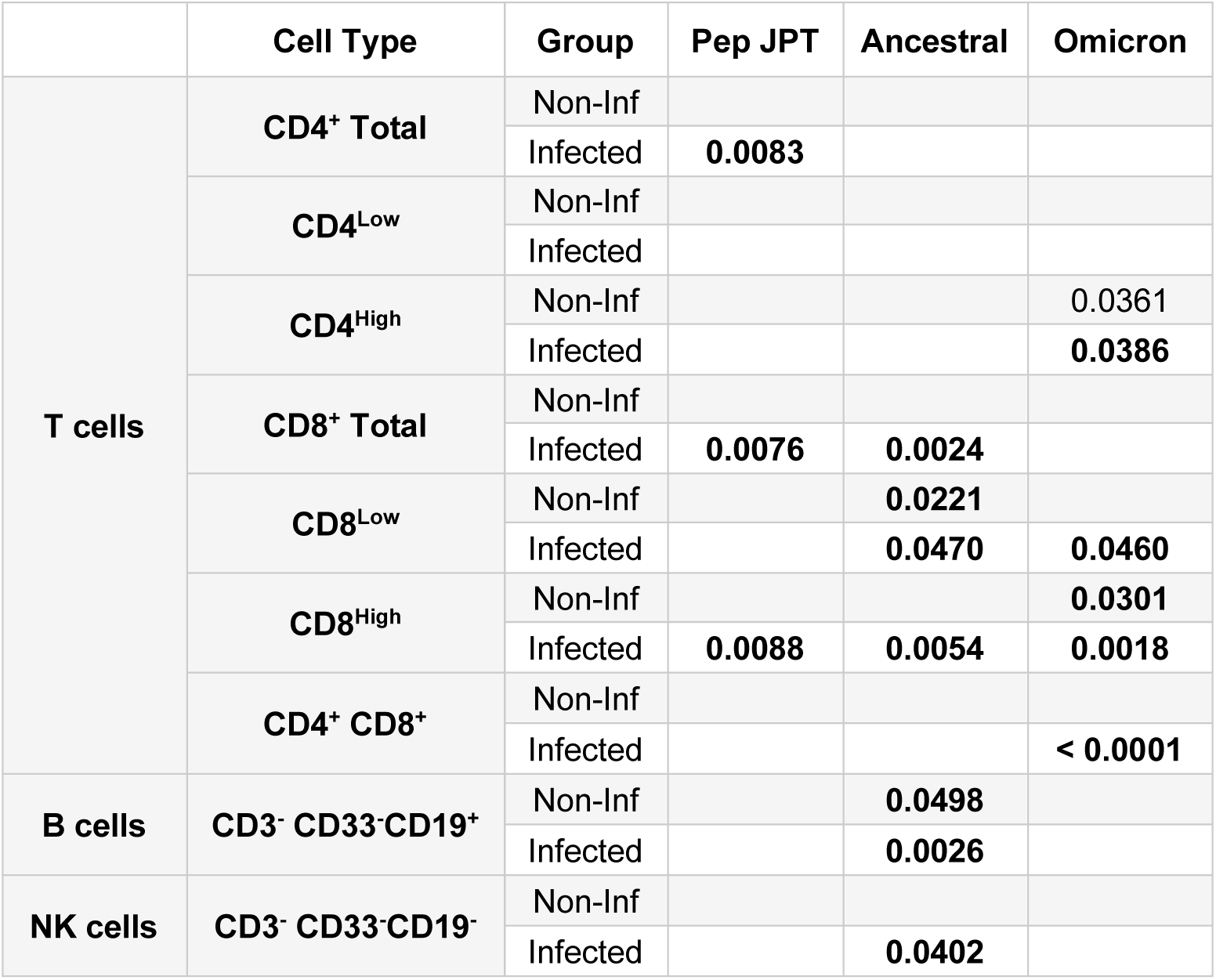
Proliferation assay shows increased response following SARS-CoV-2 infection. Comparison between both visits for the non-infected and infected groups separately. Values in bold represent an increase in the proliferation index at V3, while plain values represent a decrease at V3. Data were analized with Wilcoxon two-tailed paired t test.

The ability of the different SARS-CoV-2 strains and the spike-peptide mix to stimulate TNF and IFN-γ production in activated PBMCs was analyzed. TNF (**Fig. 9A-B**) and IFN-γ (**Fig. 9C-D**) expression were significantly stronger in peptide-stimulated PBMCs from infected compared to non-infected participants prior to and after infection. Overall, these results showed that PBMCs from infected participants, produced more cytokines in response to proliferative antigens, which contrasts with their reduced innate response (**Fig. 6**).

**Figure 9.**
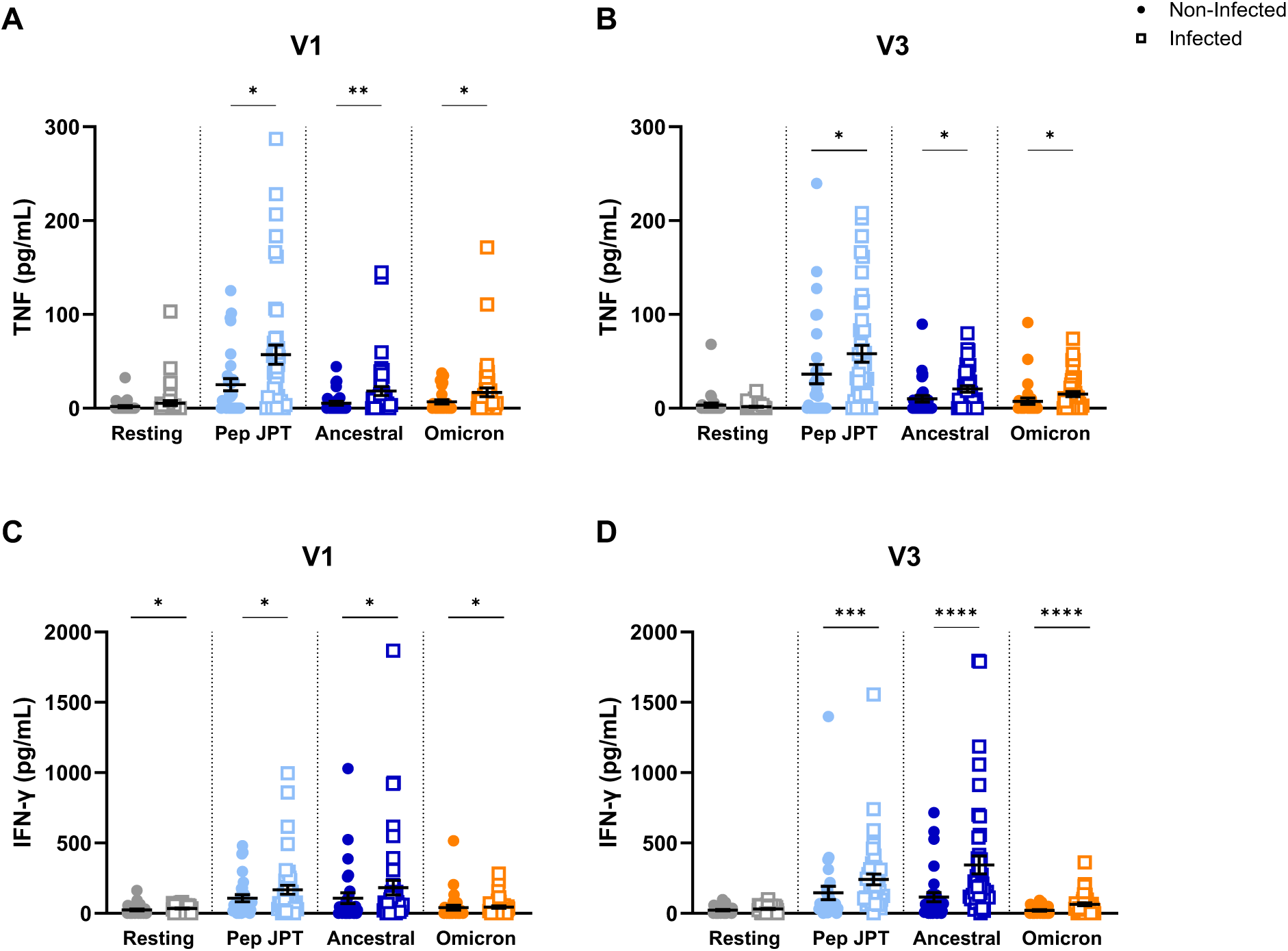
Infected group produce more TNF and IFN-γ following PBMC stimulation than non-infected. Levels of TNF and IFN-γ after PBMC stimulation under different conditions before (V1) and during (V3) Omicron predominance. TNF responses at **A)** V1 and **B)** V3, in non-infected (n=31) and infected individuals (n=44), and IFN-γ responses at **C)** V1 and **D)** V3. Each dot represents a single participant. Data are shown as mean ± SEM and were analized with Mann-Whitney two-tailed unpaired t test. * p < 0.05; ** p < 0.01; *** p < 0.001; **** p < 0.0001. Resting: no stimulation; Pep JPT: pool of peptides derived from the SARS-CoV-2 Spike glycoprotein; Ancestral: inactivated ancestral strain of SARS-CoV-2; Omicron: inactivated sublineage BA.1 Omicron variant of SARS-CoV-2.

To define immune correlates of susceptibility to Omicron BA.1 infection, we performed receiver operating characteristic (ROC) analyses on all immune parameters measured at baseline (V1), prior to viral exposure. ROC analyses were used to quantify the discriminatory capacity of individual immune features and identify signatures associated with subsequent infection. Significant parameters, including area under the curve (AUC), confidence intervals, and p-values, are summarized in **Table 3**. The strongest discriminatory power was observed for the proliferative capacity of CD4^+^ and CD8^+^ T cell subsets, particularly CD4^High^ and CD8^High^ populations, underscoring the importance of cellular immune fitness in protection against infection. Additional discriminating features included anti-N IgG3 levels, circulating I-TAC concentrations, frequencies of CD33^+^, CD4^+^, CD8^Low^, and CD19^+^ cells following 7-day stimulation, as well as TNF and IFN-γ production by virally stimulated PBMCs. We further applied ROC analyses to samples collected at V3 to distinguish individuals who experienced Omicron infection from those who remained uninfected (**Table S5**). Representative ROC curves for serological markers at V1 and V3 are shown in **Fig. 10** and reveal enhanced group separation following infection. Notably, anti-N IgG3 remained significantly discriminatory at both time points (**Fig. 10F**), identifying this parameter as a robust biomarker associated with both baseline susceptibility and the immunological imprint left by infection.

**Figure 10.**
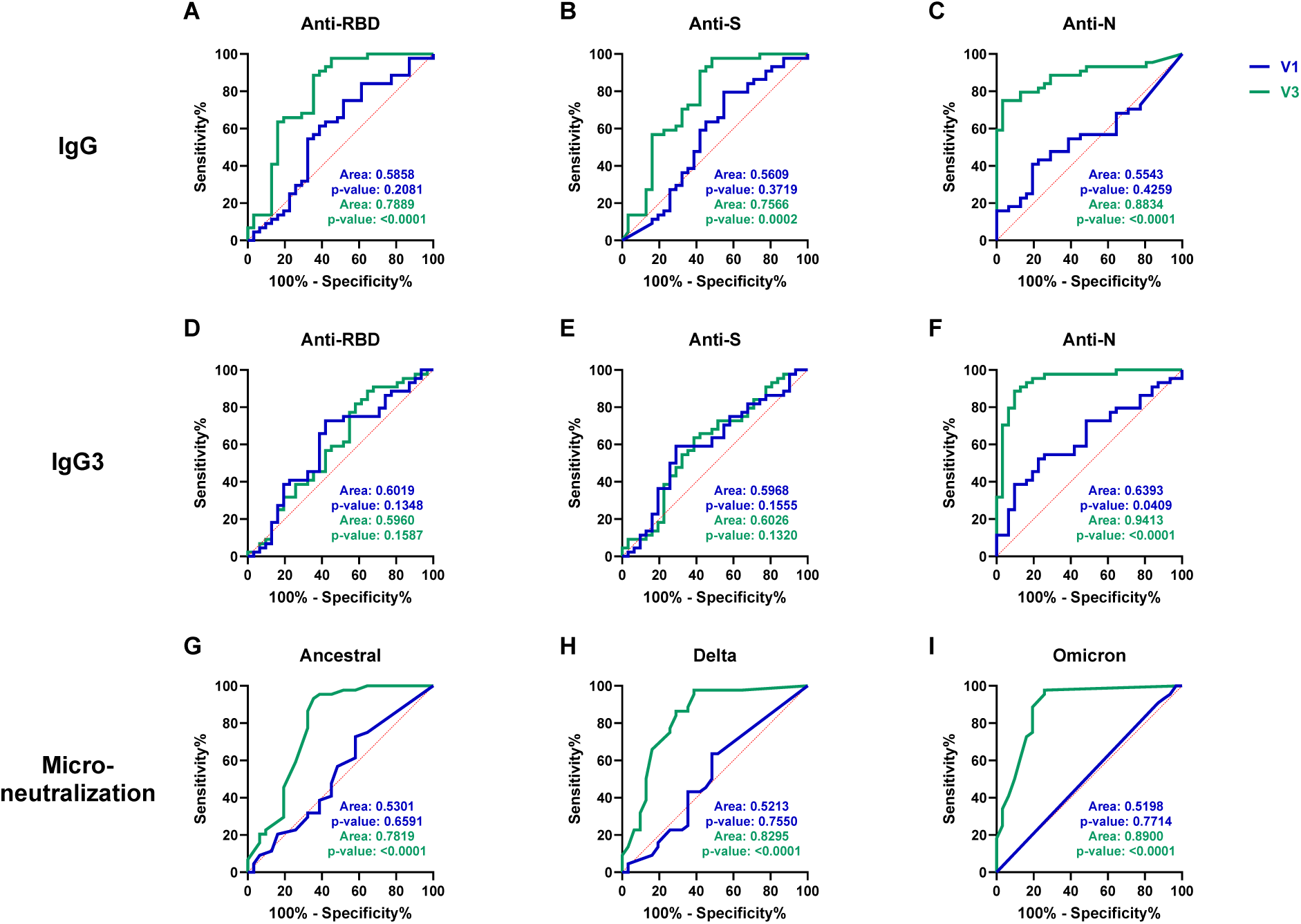
Serological parameters successfully discriminate between groups after infection. ROC curves of humoral immunity parameters to discriminate between non-infected and infected groups at V1 (blue line) and V3 (green line). IgG results for **A)** anti-RBD, **B)** anti-S, and **C)** anti-N. Results for IgG3 **D)** anti-RBD, **E)** anti-S, and **F)** anti-N. Microneutralization results against the **G)** ancestral, **H)** Delta, and **I)** Omicron BA.1 SARS-CoV-2 strains.

**Table 3.** Impairment of several immune parameters prior to infection (V1) predict subsequent Omicron BA.1 infection. Receiver operating characteristic analysis was performed with GraphPad Prism 10.5.0, with confidence interval calculated with the Wilson/Brown method. AUC: area under the curve.

| Category | Variable | Condition | AUC | Std. Error | 95% C.I. | P-value |
| --- | --- | --- | --- | --- | --- | --- |
| Antibody | IgG3 | Anti-N | 0.6393 | 0.06394 | 0.5140 - 0.7646 | 0.0409 |
| Cytokine | I-TAC | - | 0.6433 | 0.06416 | 0.5176 - 0.7691 | 0.0354 |
| Phenotype | CD33<sup>+</sup> | Resting | 0.6621 | 0.06372 | 0.5372 - 0.7870 | 0.0185 |
|  |  | Pep JPT | 0.6595 | 0.06635 | 0.5294 - 0.7895 | 0.0205 |
|  |  | Ancestral | 0.6841 | 0.06195 | 0.5627 - 0.8055 | 0.0075 |
|  |  | Omicron | 0.7356 | 0.05963 | 0.6187 - 0.8525 | 0.0006 |
|  | CD4<sup>+</sup> | Resting | 0.642 | 0.06575 | 0.5132 - 0.7709 | 0.039 |
|  |  | Ancestral | 0.672 | 0.06285 | 0.5488 - 0.7952 | 0.0124 |
|  |  | Omicron | 0.6549 | 0.06586 | 0.5258 - 0.7840 | 0.0244 |
|  | CD8<sup>Low</sup> | Pep JPT | 0.6803 | 0.0644 | 0.5541 - 0.8065 | 0.0088 |
|  | CD19<sup>+</sup> | Resting | 0.6833 | 0.06448 | 0.5570 - 0.8097 | 0.0077 |
|  |  | Pep JPT | 0.672 | 0.06634 | 0.5419 - 0.8020 | 0.0124 |
|  |  | Ancestral | 0.6701 | 0.06555 | 0.5416 - 0.7985 | 0.0134 |
|  |  | Omicron | 0.6712 | 0.06503 | 0.5438 - 0.7987 | 0.0128 |
| Proliferation index | CD4<sup>+</sup> | Pep JPT | 0.7247 | 0.06221 | 0.6027 - 0.8466 | 0.0022 |
|  |  | Ancestral | 0.7394 | 0.06099 | 0.6198 - 0.8589 | 0.001 |
|  |  | Omicron | 0.7693 | 0.05948 | 0.6527 - 0.8859 | 0.0002 |
|  | CD4<sup>High</sup> | Pep JPT | 0.6707 | 0.0671 | 0.5392 - 0.8022 | 0.0205 |
|  |  | Ancestral | 0.7332 | 0.06264 | 0.6105 - 0.8560 | 0.0015 |
|  |  | Omicron | 0.7237 | 0.06442 | 0.5974 - 0.8500 | 0.0023 |
|  | CD8<sup>+</sup> | Pep JPT | 0.7202 | 0.06322 | 0.5963 - 0.8441 | 0.0028 |
|  |  | Ancestral | 0.7477 | 0.06076 | 0.6286 - 0.8667 | 0.0006 |
|  |  | Omicron | 0.7722 | 0.05828 | 0.6580 - 0.8864 | 0.0002 |
|  | CD8<sup>High</sup> | Pep JPT | 0.7923 | 0.05785 | 0.6789 - 0.9057 | < 0.0001 |
|  |  | Ancestral | 0.8224 | 0.05381 | 0.7169 - 0.9279 | < 0.0001 |
|  |  | Omicron | 0.7746 | 0.05915 | 0.6587 - 0.8905 | 0.0002 |
| Proliferation assay ELISA | TNF | Pep JPT | 0.667 | 0.06377 | 0.5420 - 0.7920 | 0.0152 |
|  |  | Ancestral | 0.6712 | 0.06272 | 0.5483 - 0.7941 | 0.012 |
|  |  | Omicron | 0.6477 | 0.06572 | 0.5189 - 0.7765 | 0.0302 |
| | IFNγ | Pep JPT | 0.6364 | 0.06781 | 0.5035 - 0.7693 | 0.0475 |
|  |  | Ancestral | 0.6584 | 0.06703 | 0.5270 - 0.7897 | 0.0201 |
|  |  | Omicron | 0.6466 | 0.06642 | 0.5164 - 0.7768 | 0.0314 |

**Figure 11.**
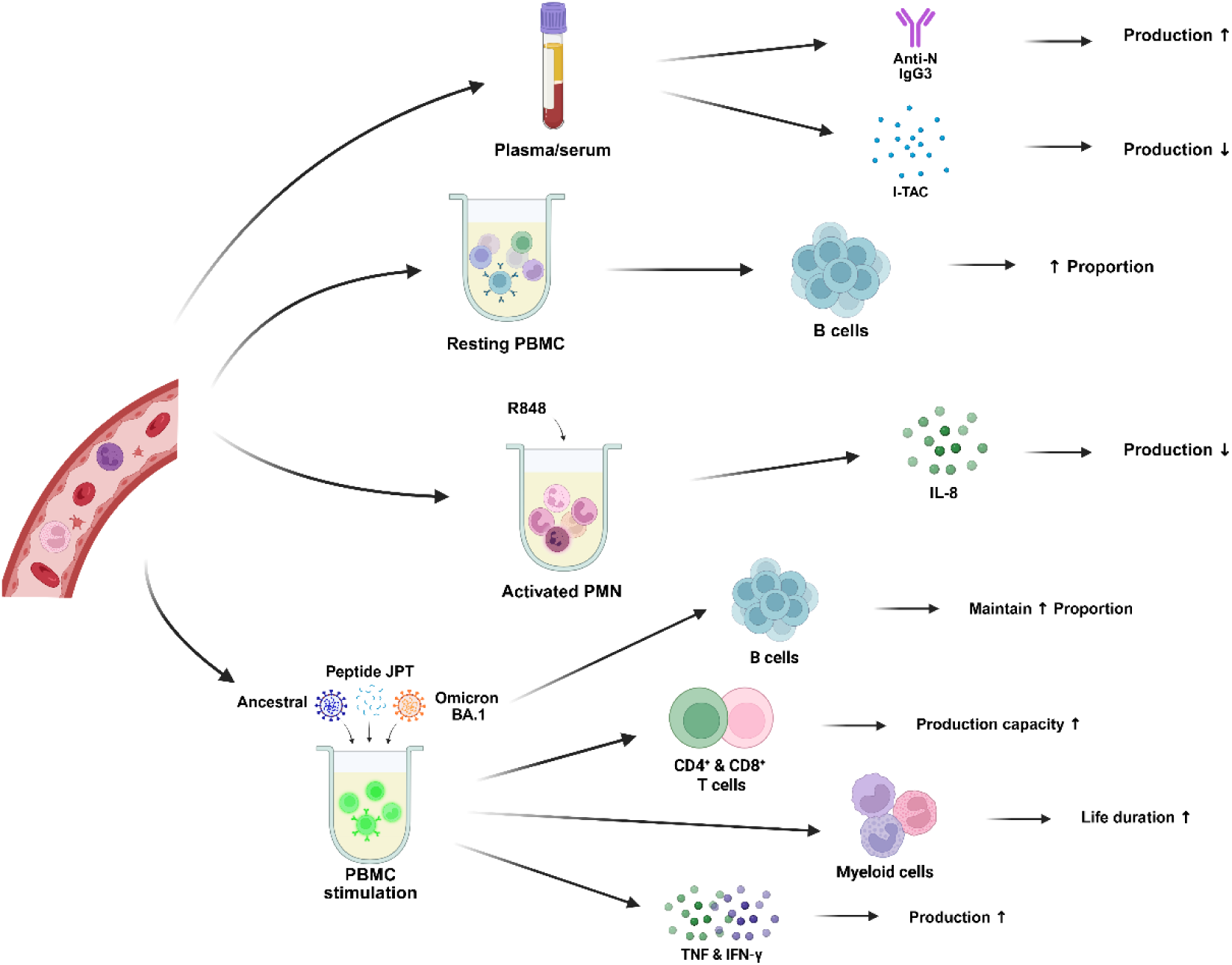
Overall representation of significant differences that increase Omicron infection susceptibility. In fully vaccinated participants before Omicron infection, those who will become infected, when compared to controls without infection, were found to have less I-TAC and more anti-N IgG3 antibodies in their plasma/serum prior being infected. Those same participants, when their PBMCs were stimulated with inactivated viruses or peptides, had lower capacity of CD4^+^ and CD8^+^ T cells proliferation, while their myeloid cells were able to survive longer, and they maintained a higher proportion of B cells. Finally, their PBMCs also produced more TNF and IFN-γ following stimulation. Figure was created with Biorender.

## DISCUSSION

The emergence of antigenically divergent SARS-CoV-2 variants has challenged current paradigms of immune protection and highlighted important gaps in our understanding of the mechanisms governing susceptibility to infection. Although neutralizing antibodies remain a key mediator against symptomatic disease [34, 35], growing evidence indicates that protection against infection is a complex sequence of coordinated interactions between the innate and adaptive immune compartments rather than the consequence of any single immune parameter [37, 39, 78]. By integrating longitudinal analyses of humoral, cellular, and innate immune responses in vaccinated individuals exposed to the first Omicron BA.1 wave, our study identified a multidimensional immune state associated with susceptibility to breakthrough infection.

Consistent with previous studies, infection resulted in enhanced antibody titers, increased serum neutralizing activity, and expansion of IgG3 responses, all characteristic features of hybrid immunity [1, 79]. The increased IgG3 response is particularly noteworthy because of IgG3’s superior Fc receptor engagement, complement activation, and antiviral effector capacity compared to other IgG subclasses [80, 81]. However, these quantitative increases do not fully explain the increased susceptibility infection. Instead, our findings indicate that qualitative and organizational features of immune responses play a critical role in determining susceptibility.

A particularly striking observation was the differential relationship between antibody binding and antiviral function according to infection susceptibility. Before viral exposure, neutralizing activity closely tracked anti-S and anti-RBD antibody levels in individuals who later became infected, suggesting that neutralization was largely driven by antibody abundance. Following infection, however, these relationships dissociated despite the marked expansion of humoral responses. In contrast, individuals who remained uninfected exhibited a persistent coupling between Spike-specific antibody levels and neutralizing capacity. This divergence suggests that protection may depend less on the magnitude of the antibody response than on the functional coordination linking antigen recognition to antiviral activity. Such coordination likely reflects qualitative properties of the antibody repertoire, including epitope selection, affinity maturation, breadth, and Fc-dependent functions, all of which have emerged as key determinants of immunity to highly evolved SARS-CoV-2 variants [2, 79, 82].

Supporting this concept, anti-N IgG3 antibodies emerged as one of the strongest baseline discriminators of subsequent infection in ROC analyses. Because nucleocapsid-directed antibodies are generally non-neutralizing, their predictive value is unlikely to reflect direct antiviral protection. Rather, elevated anti-N IgG3 responses may represent a marker of prior antigenic exposure and immune imprinting, reflecting distinct trajectories of immune memory formation [1, 2]. These findings suggest that immune history may shape susceptibility to future infection not only through the magnitude of recall responses but also through their functional organization.

Importantly, susceptibility was also associated with evidence of impaired innate immune fitness. CXCL11 was the only soluble mediator significantly reduced before infection and was among the strongest ROC-defined predictors of susceptibility. As a key IFN-γ-inducible ligand of CXCR3, CXCL11 orchestrates recruitment of antiviral effector populations including Th1 cells, cytotoxic CD8^+^ T cells, and NK cells [83]. Reduced circulating CXCL11 levels therefore suggest diminished interferon-driven immune surveillance and may indicate suboptimal antiviral priming before viral exposure. The convergence of biological function and predictive performance highlights CXCL11 as a mechanistically relevant biomarker linking interferon responsiveness to protection against infection.

Consistent with this interpretation, individuals who subsequently became infected also displayed evidence of impaired neutrophil responsiveness. Reduced IL-8 production following TLR7/8 stimulation was already apparent before infection and became more pronounced after Omicron exposure. TLR8 activation is a critical pathway regulating neutrophil antimicrobial functions, including cytokine secretion, degranulation, phagocytosis, oxidative burst, and NET formation [47, 84, 85]. Given the importance of neutrophils as first responders during viral infection, diminished IL-8 production may reflect a reduced capacity to initiate efficient innate immune recruitment and antiviral defense. Increasing evidence also indicates that neutrophils actively regulate adaptive immunity through interactions with antigen-presenting cells and T lymphocytes [86, 87], raising the possibility that impaired innate responsiveness contributes directly to the T-cell defects observed in susceptible individuals.

Indeed, one of the most consistent features associated with infection was reduced antigen-specific T-cell fitness. Long-term proliferation assays revealed diminished baseline proliferative capacity among both CD4^+^ and CD8^+^ T-cell populations in individuals who subsequently became infected. These findings are particularly relevant given the well-established role of cellular immunity in controlling SARS-CoV-2 infection and limiting disease progression, especially when viral variants evade antibody recognition [37, 40]. Notably, infection partially restored proliferative responses, consistent with the enhancing effects of hybrid immunity on T-cell memory. Nevertheless, the presence of reduced T-cell fitness prior to exposure suggests that impaired cellular responsiveness may represent a key determinant of susceptibility, even in the presence of apparently adequate humoral immunity.

Beyond functional alterations, susceptible individuals displayed a broader reconfiguration of immune organization characterized by expansion of myeloid populations and B cells accompanied by reduced CD4^+^ T-cell representation. This imbalance persisted following stimulation and remained evident after infection, suggesting the existence of a durable immune architecture associated with vulnerability. Consistent with this notion, Omicron infection induced a broad suppression of cytokine and chemokine networks while simultaneously increasing calprotectin levels, a marker of myeloid activation and inflammation. Together, these findings support a model in which breakthrough infection is associated with incomplete restoration of immune homeostasis and persistent myeloid skewing.

Collectively, our data support a paradigm in which susceptibility to SARS-CoV-2 infection is not driven by deficiencies in isolated immune pathways but rather by a pre-existing state of functional immune discoordination involving humoral, innate, and cellular compartments. The convergence of impaired interferon-associated signaling, reduced neutrophil responsiveness, diminished T-cell fitness, altered humoral coordination, and persistent myeloid inflammation identifies a multidimensional immune vulnerability that precedes infection despite robust vaccine-induced immunity. More broadly, these findings argue that protective immunity is best understood as the emergent property of an integrated immune network and establish immune coordination as a central determinant of resilience against antigenically evolving viral pathogens.

### Limitations of the Study

This study has several limitations that should be considered. First, the sample size was modest, which may have limited statistical power for some analyses and reduced our ability to detect more subtle immune differences between groups. Second, immune profiling was performed primarily using peripheral blood samples, which may not fully reflect immune responses occurring at mucosal sites, the primary portals of SARS-CoV-2 infection. In addition, the proliferation assay did not include markers allowing detailed characterization of T-cell differentiation states, such as naïve, memory, effector, or exhausted subsets. As a result, the cellular mechanisms underlying the observed differences in proliferative capacity could not be fully resolved. Similarly, the flow cytometry panel lacked specific markers for definitive identification of NK cells, requiring their characterization through an exclusion-based gating strategy. Furthermore, functional analyses were performed using bulk PBMC cultures, which do not fully reproduce the complexity of in vivo immune interactions and exclude the contribution of granulocytes and tissue-resident immune populations. Although the use of highly purified inactivated viral preparations provided a physiologically relevant stimulation model, cellular responses measured ex vivo may not fully reflect antiviral responses occurring during natural infection. Finally, despite the longitudinal design and the availability of pre-infection samples, the observational nature of the study does not allow causal conclusions regarding whether the identified immune features directly mediate susceptibility to Omicron infection. Rather, these parameters should be considered correlates of susceptibility that warrant further mechanistic investigation.

## Resource Availability

The study protocol, results and informed consent documents will be made available to researchers upon request from the corresponding author. Researchers will be asked to complete a concept sheet for their proposed analyses to be reviewed, and the investigators will consider the overlap of the proposed project with active or planned analyses and the appropriateness of the study data for the proposed analysis.

## Acknowledgements

The authors thank the study coordinator, Isabelle Chabot and the nurses of the Recherche Clinique en infectiologie *(“Centre de recherche du CHU de Québec-Université Laval”)* for their involvement in this study. We are very grateful to the study participants, without whom this study would not have been feasible. Thanks to the National Microbiology Laboratory (NML), Public Health Canada, for providing the SARS-CoV-2 isolate (SARS-CoV-2 VOC B1.1.529 Omicron sub- lineage BA.1) used in this study. The authors thank Dr. Nicolas Chomont for the qPCR protocol, Dr. Martin Pelletier for access to the qPCR platform, Dr Yann Breton, Dr Annie Gravel and Audrey Edmond for helping with Luminex and cytometer standardization. The authors thank the *“Plateforme de Bio-fabrication de Vaccins Québec/Centre de Recherche du CHU de Québec-Université Laval”* for the MESO^®^ QuickPlex SQ 120MM service.

This project is being supported by funding from the Public Health Agency of Canada, through the Vaccine Surveillance Reference group and the COVID-19 Immunity Task Force (grant number: 2021-HQ-000134) to DB, MB, CG, JFM, JFM, and ST. JB and WBB are the recipients of the Desjardins scholarship from the Fondation du CHU de Québec, and the recruitment scholarship from the AIDS Research Fund of Université Laval. WWB is the recipient of the leadership and sustainable development scholarship, and the Fonds de recherche du Québec-Santé (FRQ-S) doctoral training scholarship. MB was supported by the Sentinel North Research Chair at Université Laval, funded by the Canada First Research Excellence Fund and by the Canada Research Chair. This work was also supported by the Fonds de recherche du Québec (FRQ) through the research centre grant for the CHU de Québec-Université Laval Research Center (reference: 30641).

## Author contributions

HJ: Conceptualization, Data curation, Formal analysis, Investigation, Methodology, Writing – original draft, Writing – review & editing. WWB: Data curation, Formal analysis, Investigation, Methodology, Writing – review & editing. IB: Investigation, Methodology, Writing – review & editing. BG: Formal analysis, Investigation, Methodology, Writing – review & editing. JB: Investigation, Writing – review & editing. AP: Investigation, Writing – review & editing. KDC: Investigation, Writing – review & editing.HR: Investigation, Writing – review & editing. MT: Writing – review & editing, Data curation, Formal analysis, Investigation, Methodology, Validation. KS: Methodology, Writing – review & editing. M-AL: Investigation, Resources, Writing – review & editing. J-FM: Funding acquisition, Validation, Writing – review & editing. JNP: Funding acquisition, Resources, Writing – review & editing. NB: Formal analysis, Writing – review & editing. DB: Funding acquisition, Project administration, Writing – review & editing. ST: Funding acquisition, Project administration, Resources, Writing – review & editing. MB: Funding acquisition, Investigation, Resources, Writing – review & editing. CG: Conceptualization, Formal analysis, Funding acquisition, Investigation, Methodology, Project administration, Resources, Supervision, Validation, Writing – original draft, Writing – review & editing.

## Declaration of generative AI and AI-assisted technologies in the writing process

During the preparation of this work, the authors used Biorender’s AI tool in order to keep the same legend between the figures. After using this tool/service, the authors reviewed and edited the content as needed and take full responsibility for the content of the published article.

## Ethical Statement

The study was approved by the “Centre de Recherche du CHU de Québec-Université Laval” (Québec, QC, Canada) ethics review boards (CER- 2021-5744) and conducted according to the guidelines of the Declaration of Helsinki. All participants provided written informed consent prior to enrollment. All experiments with SARS-CoV-2 and participant’ samples were performed in the Containment Level 2 and 3 (CL2, CL3) laboratories at the “CHU de Québec-Université Laval”.

## METHODS

### Study Population

A cohort of 304 retail workers in the Québec City metropolitan area was originally recruited early in the pandemic to study SARS-CoV-2 infection rate and vaccination uptake as described previously [73, 88–92]. Briefly, eligibility criteria included the following: age ≥ 18 years; having a public-facing role in work-related activities; and having no history of hospitalization due to COVID- 19 (69). Study participants were followed every twelve weeks over five visits, with data collected on socio-demographic and clinical characteristics, COVID-19 vaccination, SARS-CoV-2 symptoms and testing. Blood was drawn for measurement of SARS-CoV-2 antibody titers, serum neutralization assays, and cellular immune responses. SARS-CoV-2 infection was defined as having reported at least one positive nasopharyngeal test (PCR or antigen detection) or having serological evidence of SARS-CoV-2 infection (co-positivity of anti-spike and anti-nucleocapsid IgG antibodies). The present study compares two subgroups of participants who had no SARS-CoV-2 infection at their first visit (V1), and who either became infected or not by the third visit (V3). V1 took place before Omicron predominance, while V3 occurred twenty-four weeks later during Omicron predominance. The immune response to SARS-CoV-2 in the infected group was compared to the immune response in a non-infected group (**Table 4**). As we have previously shown that innate responses differed across seasons [92], we selected participants whose visits occurred during the same seasons (see Flow Chart **Fig. 1S**).

**Table 4.** Participant description at entry (V1) Median was analyzed with the Mann-Whitney test, the rest of the data was analyzed with Fisher’s exact test. IQR: interquartile range. ^1^See **Table S1. SARS-CoV-2 Exposition Levels**

| Variables n (%) | All participants<br>n=75 | Non-infected<br>n= 31 | Infected<br>n = 44 | P-Value |
| --- | --- | --- | --- | --- |
| **Age** |  |  |  |  |
| Median (IQR) | 34.0 (25.0-48.0) | 34.0 (22.0-51.0) | 33.0 (25.3-47.8) | 0.5980 |
| [18, 25] | 22 (29.3) | 11 (35.5) | 11 (25) | 0.4544 |
| [26, 44] | 31 (41.3) | 10 (32.3) | 21 (47.7) |  |
| [45, 59] | 18 (24.0) | 9 (29.0) | 9 (20.5) |  |
| [60, +[ | 4 (5.3) | 1 (3.2) | 3 (6.8) |  |
| **Birth Sex** |  |  |  |  |
| Female | 48 (64.0) | 18 (58.1) | 30 (68.1) | 0.4651 |
| Male | 27 (36.0) | 13 (41.9) | 14 (31.8) |  |
| **Level of exposure to SARS-CoV-2<sup>1</sup>** |  |  |  |  |
| Low | 32 (42.7) | 16 (51.6) | 16 (36.4) | 0.2382 |
| Medium | 43 (57.3) | 15 (48.4) | 28 (63.6) |  |
| **COVID-19 vaccine** |  |  |  |  |
| **Doses received** |  |  |  |  |
| 0 dose | 5 (6.7) | 3 (9.7) | 2 (4.5) | 0.3245 |
| 1 dose | 8 (10.7) | 5 (16.1) | 3 (6.8) |  |
| 2 doses | 61 (81.3) | 23 (74.2) | 38 (86.4) |  |
| 3 doses | 1 (1.3) | 0 (0.0) | 1 (2.3) |  |
| **Type of vaccine received** |  |  |  |  |
| **Dose 1** |  |  |  |  |
| Pfizer | 38 (54.3) | 13 (46.4) | 25 (59.5) | 0.4849 |
| Moderna | 19 (27.1) | 8 (28.6) | 11 (26.2) |  |
| AstraZeneca | 13 (18.6) | 7 (25.0) | 6 (14.3) |  |
| **Dose 2** |  |  |  |  |
| Pfizer | 34 (54.8) | 12 (52.2) | 22 (56.4) | > 0.9999 |
| Moderna | 26 (41.9) | 10 (43.5) | 16 (41.0) |  |
| AstraZeneca | 2 (3.2) | 1 (4.3) | 1 (2.6) |  |
| **Dose 3** |  |  |  |  |
| Pfizer | 1 (100.0) | 0 (0.0) | 1 (100.0) |  |

### Blood Collection and PMN and PBMC Purification

Peripheral blood was collected using serum, citrate, heparin, and EDTA tubes. Samples were then transferred to the laboratory for processing within two hours. Briefly, tubes were centrifuged to collect and aliquot the serum and plasma. Plasma was further centrifuged at 3000 x *g* for 10 min to remove platelets. Serum and plasma samples were frozen at -80°C until use. The remaining blood was further processed to purify PBMCs and PMNs.

PBMCs were isolated from peripheral blood collected in EDTA-containing tubes using Lymphocyte Separation Medium (Wisent, Cat: 305-010-CL) density gradient centrifugation as described previously [92]. Cells (3 x 10^6^) were used for R848 (Resiquimod, InvivoGen, Cat: vac-r848) stimulation, and the rest was stored at -150°C in freezing medium consisting of fetal bovine serum (FBS, Sigma, Cat: F1051) with 10% cell culture-grade dimethylsulfoxyde (DMSO, Sigma, Cat: D2650) until use.

PMNs were isolated from peripheral blood collected in EDTA-containing tubes as previously described [92]. First, 2% dextran sedimentation of erythrocytes was performed, followed by Lymphocyte Separation Medium (Wisent, Cat: 305-010-CL) density gradient centrifugation. Hypotonic lysis was used to eliminate the remaining contaminating erythrocytes. Purified PMNs were then used for R848 stimulation.

### Antibody Assays

Antibody response (anti-spike (anti-S), anti-receptor binding domain (anti-RBD), anti-nucleocapsid (anti-N) antibodies as well as IgG subclasses) to SARS-CoV-2 was assessed by two enzyme-linked immunosorbent assays (ELISA), chemiluminescent [91] and colorimetric approaches.

The chemiluminescent ELISAs were developed at the Universities of Ottawa and Toronto and were calibrated against a World Health Organization reference standard [91]. Briefly, automated chemiluminescent ELISAs were performed as described using robotic liquid handlers [91]. Antigens (spike, RBD, nucleocapsid; 50 ng/well) were coated on 384-well plates overnight at 4 °C. After blocking with 3% skim milk in PBST, diluted samples and controls were added and incubated for 2 h. HRP-conjugated secondary antibodies (anti-IgG or anti-IgG3) were then added, followed by a chemiluminescent substrate. Signal was measured using a plate reader after a short incubation. For calibration, a pooled standard of 13 COVID-19-positive samples was serially diluted (12 steps). Relative concentrations (ranging from 1 to 2048) were assigned arbitrarily. Each dilution was tested in quadruplicate across three plates, with negative and blank controls included.

### In-house ELISA for antibody monitoring

The semi-automated in-house colorimetric ELISA was performed as previously described [91]. Briefly, 96-well plates were coated overnight at 4 °C with 100 µL of SARS-CoV-2 antigen. Plates were washed and blocked with 3% skim milk for 1 h at room temperature (RT), then washed again. Heat-inactivated serum samples (56 °C, 1 h) were diluted as follows: 1:15,000 for anti-S IgG, 1:200 for anti-N IgG, and 1:100 for IgM, added to the plates (100 µL), and incubated for 1 h at RT. After washing, HRP-conjugated secondary antibodies were added (anti-IgG 1:30,000; anti-IgM 1:10,000) and incubated for 1 h. Following additional washes, tetramethylbenzidine (TMB) substrate was added for 1 h, and the reaction was stopped with 2 M HCl. Absorbance was read at 450 nm. Serological evidence of SARS-CoV-2 infection was defined as co-positivity of anti-S and anti-N antibodies.

### Microneutralization Assay

Microneutralization assays, the gold standard for evaluating virus neutralizing antibodies, were performed as previously described [73]. Sera were heat-inactivated, serially diluted (1:20-1:10240) and incubated with 100 TCID₅₀ of each SARS-CoV-2 strain for 1 h at RT. The residual infectivity was assessed in Vero E6 (ancestral, Delta) or Vero TMPRSS2 (Omicron BA.1) cells. Neutralizing titers were defined as the highest serum dilution completely preventing cytopathic effect at day 4. Results are expressed as geometric mean titers (GMT). Microneutralization assays were performed in the Containment Level 3 (CL3) laboratory at the “CHU de Québec-Université Laval”.

### TLR7/8 Stimulation

PBMCs and PMNs were resuspended at 2 x 10^6^ cells/mL in high-performance TheraPEAK™ X- VIVO™-15 Serum-free Hematopoietic Cell Medium (Lonza, Cat: BEBP04-744Q). The cells were plated in round-bottom 96-well plates at 100,000 cells/well and 200,000 cells/well, and stimulated with the viral analog R848 (Resiquimod, InvivoGen, Cat: vac-r848) in triplicate with four different R848 conditions (0, 0.5, 1 and 2 μg/mL) for 24 h at 37°C in 5% CO_2_. The plates were then centrifuged 5 min at 400 *x g*, and the supernatant was transferred to a flat-bottom 96-well plate and kept at -20°C until use.

### ELISA for Immune-Inflammation Markers Monitoring

IL-8 (Cat: DY208), TNF (Cat: DY210) and IFN-γ (Cat: DY285B) concentrations were measured by ELISA following the manufacturer’s instructions (R&D Systems). Briefly, 96-well plates were coated with mouse anti-human capture antibody overnight. They were then rinsed three times with wash buffer (0.05% Tween-20 in PBS) and blocked for 1 h with blocking buffer (1% BSA in PBS, pH 7.2-7.4, 0.2 μm filtered, R&D Systems, Cat: DY995). After rinsing, samples, standards, and controls were added and incubated for 2 h at RT. Plates were then washed, and biotinylated goat (IL-8, TNF) or mouse (IFN-γ) anti-human detection antibody was added to the wells for 2 h. After washing, streptavidin-HRP solution was added and left to incubate for 20 min in the dark. Plates were washed and prepared for the detection of bound antibodies by adding the substrate solution (1:1 mixture of H_2_O_2_ and TMB (R&D Systems, Cat: DY999) for 20 min in the dark, followed by the stop solution (2N H_2_SO_4_, R&D Systems, Cat: DY994). Finally, plates were read at 450 and 540 μm with the SpectraMax 190 microplate reader (Molecular Devices, San Jose, CA, USA).

### Plasma Cytokine Multiplex Dosing

Plasma cytokines were measured using the recommended protocol for the ProcartaPlex^TM^ Human Immune Monitoring Panel 65-Plex (ThermoFisher Scientific Inc., Cat. EPX650-10065-901). Briefly, capture beads were added to the plate and washed twice with the provided wash buffer. Undiluted samples, standards, and controls were added to the plate and incubated for 2 h at RT with shaking. The plate was then washed twice and the biotinylated detection antibodies were added and incubated for 30 min with shaking. The plate was washed again and streptavidin-PE was added for 30 min with shaking. Finally, the plate was washed, reading buffer was added and left to incubate for 5 min with shaking before reading with the Bio-Rad Bio-Plex 200 system (Bio-Rad Laboratories, CA, USA). Values that were below the lower limit of detection were reported as half the lowest concentration measured for each cytokine.

### Plasma Calprotectin Measurement

Plasma calprotectin was measured by ELISA using the recommended protocol for the R-PLEX Human Calprotectin Assay (Meso Scale Discovery, Rockville, MD, USA, Cat: K151AJYR-2). All steps were performed at RT. Briefly, the capture antibody was added to 96-well plates and was incubated with shaking for 1 h. Following a washing step, the standards and diluted samples (1:100) were added to the plates and left to incubate 1 h with shaking. Plates were washed and the detection antibody was added for a 1-hour incubation with shaking. After a last wash, reading buffer was added and the plates were immediately read using the MESO QuickPlex SQ 120MM (Meso Scale Discovery).

### Production and Quantification of Highly Purified Inactivated Viruses

The method described by Darnell *et al.*, 2004 [93] for SARS-CoV-1 was used to prepare the samples. Quantification was done via total RNA extraction [94, 95]. Briefly, Vero E6 cells were infected with SARS-CoV-2 virus isolated from a clinical sample collected in March 2020 in Quebec City, Canada (SARS-CoV-2/Quebec City/21697/2020, ancestral Wuhan 1-like SARS-CoV-2), while Vero TMPRSS2 cells were infected with the Omicron sublineage BA.1 obtained from the National Microbiology Laboratory (NML), Public Health Agency of Canada. Supernatants were harvested 4 days post-infection when cytopathic effect (CPE) was around 90%, and 30 mL were transferred to 50 mL Falcon tubes to be centrifuged at 2,500 rpm for 10 min at 4°C. The supernatants were transferred to other 50 mL Falcon tubes and a 1:4000 dilution of glutaraldehyde was added to each tube. The tubes were incubated at 37°C for 3 days. An aliquot of 1 mL was saved before inactivation (no glutaraldehyde added) to use as a control. Inactivated viruses were aliquoted in cryovials (1 mL each) and saved at -80°C. The following day, Vero E6 and Vero TMPRSS2 cells were incubated with 200 µL of each SARS-CoV-2 virus before and after inactivation (passage 1). Cultures were observed for 7 days and then used to perform another passage (passage 2), similar to passage 1, observed for 7 days for CPE and used to perform a third passage, which was also observed for 7 days. In all passages, CPE in the non-inactivated virus was observed from day 2. However, no CPE was detected after 7 days of incubation in any of the passages with inactivated SARS-CoV-2. Frozen production supernatants were thawed at RT and immediately ultracentrifuged at 100,000 x *g* for 60 min at 4°C in 20 mL tubes (Seton Scientific, Cat: 5082) using a fixed-angle titanium rotor T-1250 (ThermoFisher). The remaining supernatant was ultracentrifuged again under the same conditions in new 20 mL tubes. The virus pellets from both ultracentrifugations were resuspended and combined in 0.22 µm filtered PBS to obtain a 10X concentrate of the original production supernatant. An aliquot of the viral preparation was diluted in Trizol LS (ThermoFisher, Cat: 10296028) for viral RNA quantification.

### SARS-CoV-2 Quantification

Total RNA was extracted using the phenol/chloroform method and diluted in 60 µL of 1X TE buffer [96]. A 15 µL RT-qPCR reaction contained 5 µL of RNA, 3.75 µL TaqPath 1-Step Multiplex Master Mix (ThermoFisher, Cat: A28526), 1.88 µL of E gene 8x primer/probe set, 1.88 µL of N gene 8x primer/probe set, and 2.5 µL of PCR clean H_2_O [94, 95]. All primers and probes were obtained from IDT (Integrated DNA Technologies) (**Table S6**). Thermal cycling was performed at 53°C for 10 min for reverse transcription, and 95°C for 2 min for reverse transcriptase inactivation, followed by 45 cycles at 95°C for 3 seconds, and 60°C for 30 seconds for cDNA amplification. Reactions were carried in white 96-well PCR plates (Bio-Rad, Cat: HSP9601) in a CFX Connect Real-Time PCR Detection System (Bio-Rad). Quantification was performed for the initial viral preparation, the 1^st^ and 2^nd^ supernatants and the final pellet (**Fig. S11A, C, E**).

### Dynamic Light Scattering (DLS) Analysis

Inactivated viral samples were analyzed in spectrophotometer cuvettes using a Zetasizer Nano ZS (Malvern Instruments). Hydrodynamic diameter measurements were done in duplicate at RT for the initial viral preparation, the 1^st^ and 2^nd^ supernatants and the final pellet (**Fig. S11B, D, F**).

### Carboxyfluorescein Succinimidyl Ester (CFSE) Cell Proliferation Assay

PBMCs were thawed as described in appendix 1 and resuspended in RPMI (Wisent, Cat: 350- 000-CL) with 2% FBS (Sigma, Cat: F1051) containing 50 U/mL benzonase (Millipore Sigma, Cat: 1.03773.1010) or 20 µg/mL DNase (Millipore Sigma, Cat: DN25) and left to incubate at 37°C + 5% CO_2_ for 1 h. The medium was removed and cells were suspended at 1 x 10^7^ cells/mL in HBSS 1X 2mM EDTA. Eight million cells per mL were stained with 0.5 µM CFSE (Invitrogen, Cat: C34554) in HBSS 1X for 8 min. Cells were centrifuged, medium was removed, and cells were resuspended in X-VIVO^TM^ 15 medium for hematopoietic cells (Lonza, Cat: BEBP04-744Q) at 4 x 10^6^ cells/mL, and left to incubate at 37°C + 5% CO_2_ for 30 min along with unstained cells. A fraction of stained and unstained cells was reserved for flow cytometry analysis and phenotype determination at day 0. Medium was removed and cells were resuspended in X-VIVO^TM^ 15 medium at 4 x 10^6^ cells/mL. They were plated at 2 x 10^6^ cells/mL in round bottom 96-well plates. Cells were activated with either the PepMix SARS-CoV-2 (JPT innovative peptide solution, Cat: PM-WCPV-S-2; 0.06 nmol/mL), the ultrapurified attenuated ancestral virus (4.7 x 10^6^ SARS-CoV- 2 RNA copies/mL) or the ultrapurified attenuated Omicron BA.1 virus (3.3 x 10^6^ SARS-CoV-2 RNA copies/mL or 2.0 x 10^6^ SARS-CoV-2 RNA copies/mL) in duplicates. Unstimulated cells were also plated for a control resting condition. The cells were left to incubate at 37°C + 5% CO_2_ for 7 days. On day 7, plates were centrifuged 5 min at 400 x *g*. Supernatants were transferred to flat-bottom 96-well plates for subsequent ELISA analysis, and cells were stained for flow cytometry analysis (**Figure S12**).

### Staining Protocol

Cells were resuspended in HBSS 1X with 2 mM EDTA (FACS buffer) and transferred in V-bottom 96-well plates by combining duplicates/triplicates. A fraction of CFSE-stained and non-stained cells from each donor were combined to form CFSE and non-CFSE pools respectively, and were distributed in wells for controls, compensations, and FMOs at day 0. Half of the cells in a well from the non-CFSE pool was heated at 56°C for 1 min to obtain dead cells. Plates were centrifuged 5 min at 400 x *g* and cells were washed with FACS buffer and centrifuged again. Stained cells were resuspended in 0.125 μg/mL FVS450 (BD Bioscience, Cat: 562247) in FACS buffer, while unstained cells were resuspended in FACS buffer for 15 min. FACS buffer with 1% FBS (Sigma, Cat: F1051) was added, and plates were centrifuged 5 min at 400 x *g*. Supernatants were removed and Fc block solution (FACS buffer with 2% FBS, human Fc block (BD Bioscience, Cat: 564220), and mouse serum) was added to resuspend cells for 10 min. The antibody panel (**Table S7**) was added and left to incubate 30 min in the dark. Cells were then washed with FACS buffer + 1% FBS and centrifuged 400 x *g* before being fixated with a 4% PFA solution (Fisher, Cat: A11313) for 20 min. FACS buffer + 1% FBS was added before centrifuging and resuspending cells in FACS buffer, and transferring them in 5 mL polystyrene test tubes. Rainbow beads (Spherotech, Cat: RCP-30-5A) were used to calibrate the cytometer before the acquisition of every experiment (**Fig. S13**).

### Manual Phenotype Gating

First, the PBMCs were identified based on the FSC and SSC parameters. Single cells were selected by analyzing FSC-H versus FSC-A to remove doublets. Live cells were then selected using FVS450. Myeloid cells were identified based on the expression of CD33, in which CD4^+^ and CD8^+^ subpopulations were gated. T (CD3^+^) cells were gated from the CD33^-^ population, while B (CD19^+^) cells were gated on the CD33^-^ CD3^-^ population generated by a Boolean gate. CD3^+^ T cells were analyzed to gate CD4^+^ helper, CD8^+^ cytotoxic, double negative, and double positive T cells. Subsets of CD4^+^ and CD8^+^ T cells were investigated following their fluorescence intensity, and separated between low and high expression. Remaining cells (CD33^-^ CD3^-^ CD19^-^) were considered NK cells (**Fig. S14**). While cell phenotypes were identified, CFSE expression was gated in each population comparatively to day 0 cells in order to extract proliferation data from the FlowJo v10.10.0 software (BD Bioscience, Ashland, OR, USA) algorithm.

### Statistical Analysis

All statistical analyses were performed with GraphPad Prism 10.5.0 (GraphPad Inc, San Diego, CA, USA). Participant demographic and clinical characteristics were presented as a proportion or median with an interquartile range (IQR) and tabulated. Unpaired t tests were performed for continuous variable comparison, and Fisher’s exact test was used for categorical variables. For the rest of our data, initial test of normality and log normality indicated they did not fit both distributions; therefore, all values were analyzed using nonparametric tests. Lastly, to identify immune parameters associated with susceptibility to infection, all baseline (V1) data were analyzed using receiver operating characteristic (ROC) curve analysis to discriminate between non-infected and infected individuals and to estimate the probability of infection with the Omicron BA.1 variant. For each parameter, the area under the curve (AUC) was calculated as a measure of discriminatory performance, with AUC values interpreted as follows: 0.5, no discrimination; 0.6- 0.7, poor; 0.7-0.8, acceptable; 0.8-0.9, excellent; and >0.9, outstanding discrimination. Optimal thresholds were determined using Youden’s index to maximize the combined sensitivity and specificity of each marker. To account for multiple comparisons across the panel of immune parameters, p-values were adjusted using the false discovery rate (FDR) method (Benjamini– Hochberg correction), and only parameters remaining significant after correction were retained. Confidence intervals for proportions were calculated using the Wilson/Brown method. All statistical analyses were performed using standard statistical software, and results were systematically tabulated. All tests were two-sided and p-values < 0.05 were considered statistically significant.

### Supplemental Information

Document S1. Figures S1-S14 and tables S1-S6

