## Supplementaldata for "A Multidimensional Immune Signature Predicts Susceptibility to Omicron Infection in Vaccinated Individuals"

| Parameters | Answers | |
|---|---|---|
| | Yes | No |
| Participant's travel outside province since January 2020 | 2 | 0 |
| Participant's have supplemental work other than their main occupation | 1 | 0 |
| Participant's COVID-19 protective behaviours at work place: Use of mask | 0 | 2 |
| Participant's COVID-19 protective behaviours at work place: Physical distancing | 0 | 2 |
| Participant's COVID-19 protective behaviours at work place: Hand washing | 0 | 1 |
| Participant's COVID-19 protective behaviours at work place: Plexiglas | 0 | 1 |
| Participant's COVID-19 protective behaviours at work place: other additional means | 0 | 1 |
| Number of gathering of 10 people or more since March 2020 | | |
| 0 | 0 | |
| 1 to 5 | 2 | |
| 6 to 10 | 4 | |
| more than 10 | 6 | |
| Public transport used: Bus | 1 | 0 |
| Ratio of number of people in the household / number of rooms greater than 1 | 1 | 0 |
| Health worker in household | 1 | 0 |
| Number of kids under 18 years old in household | | |
| 0 | 0 | |
| 1 to 2 | 2 | |
| more than 2 | 4 | |
| Contact with suspect COVID-19 case | 3 | 0 |
| Exposure time during wave 5 | | |
| 0-25% | 1 | |
| 25-50% | 2 | |
| 50-75% | 3 | |
| 75-100% | 4 | |
| Main occupation | | |
| Bar and Restaurant | 1 | |
| Hardware store and grocery | 2 | |

**Table S1. SARS-CoV-2 exposition levels**

Evaluation criteria for the exposition level to the SARS-CoV-2 virus and the number of points attributed. Breakdown of the exposure level, following the addition of results, is the following: Low < 10 points, Medium between 10 and 20 points, and High > 20 points.

| Variables n (%) | All participants n=75 | Exposed Non-infected n= 31 | Exposed Infected n = 44 | P-Value |
|---|---|---|---|---|
| **Categories of employment or worker** | | | | |
| *Restaurant/Bar* | *39 (52.0)* | *14 (45.2)* | *25 (56.8)* | *0.4128* |
| *Grocery* | *24 (32.0)* | *10 (32.3)* | *14 (31.8)* | |
| *Hardware* | *12 (16.0)* | *7 (22.6)* | *5 (11.4)* | |
| **Education** | | | | |
| *High School or less* | *18 (24.0)* | *9 (29.0)* | *9 (20.5)* | *0.3239* |
| *Professional certification/CEGEP* | *39 (52.0)* | *13 (41.9)* | *26 (59.1)* | |
| *University degree* | *18 (24.0)* | *9 (29.0)* | *9 (20.5)* | |
| **BMI** | | | | |
| **Median (IQR)** | *25.2 (22.4-31.0)* | *25.6 (23.3-31.0)* | *25.2 (21.0-31.0)* | *0.4749* |
| *18,5 - 24,9* | *35 (46.7)* | *14 (45.2)* | *21 (47.7)* | *0.9547* |
| *25,0 - 29,9* | *18 (24.0)* | *8 (25.8)* | *10 (22.7)* | |
| *≥ 30* | *22 (29.3)* | *9 (29.0)* | *13 (29.5)* | |
| **Comorbidities** | | | | |
| **At least one comorbidity** | *19 (25.0)* | *5 (16.1)* | *14 (31.1)* | *0.1785* |
| *Hypertension* | *4 (5.3)* | *1 (3.2)* | *3 (6.7)* | |
| *Diabetes* | *6 (7.9)* | *2 (6.5)* | *4 (8.9)* | |
| *Asthma* | *7 (9.2)* | *1 (3.2)* | *6 (13.3)* | |
| *Chronic lung disease* | *1 (1.3)* | *0 (0.0)* | *1 (2.2)* | |
| *Chronic heart disease* | *1 (1.3)* | *0 (0.0)* | *1 (2.2)* | |
| *Chronic liver disease* | *1 (1.3)* | *0 (0.0)* | *1 (2.2)* | |
| *Cancer* | *1 (1.3)* | *0 (0.0)* | *1 (2.2)* | |
| *Immunodepression* | *3 (3.9)* | *1 (3.2)* | *2 (4.4)* | |
| *Chronic neurological disorder* | *2 (2.6)* | *1 (3.2)* | *1 (2.2)* | |
| **Smoker (tobacco)** | *14 (18.7)* | *6 (19.4)* | *8 (18.2)* | *> 0.9999* |
| *Occasionally* | *6 (42.9)* | *3 (50.0)* | *3 (37.5)* | |
| *Every day* | *8 (57.1)* | *3 (50.0)* | *5 (62.5)* | |
| **Vaping** | *6 (7.9)* | *2 (6.5)* | *4 (8.9)* | *> 0.9999* |
| *Occasionally* | *4 (66.7)* | *2 (100.0)* | *2 (50.0)* | |
| *Every day* | *2 (33.3)* | *0 (0.0)* | *2 (50.0)* | |

**Table S2. Additional demographic and clinical characteristics of the study subjects at entry**
Median was analyzed with the Mann-Whitney test, the rest of the data was analyzed with Fisher’s exact test. IQR: interquartile range.

| | Cell Type | Visit | Pep JPT | Ancestral | Omicron |
|---|---|---|---|---|---|
| T cells | CD4$^{+}$ Total | V1 | < 0.0001 | 0.0028 | 0.0009 |
| | | V3 | 0.0012 | < 0.0001 | 0.0013 |
| | CD4$^{Low}$ | V1 | | 0.0378 | |
| | | V3 | | 0.0067 | |
| | CD4$^{High}$ | V1 | 0.0006 | 0.0002 | 0.0082 |
| | | V3 | < 0.0001 | < 0.0001 | < 0.0001 |
| | CD8$^{+}$ Total | V1 | 0.0105 | 0.0026 | 0.0011 |
| | | V3 | < 0.0001 | 0.0026 | 0.0004 |
| | CD8$^{Low}$ | V1 | | | 0.0433 |
| | | V3 | | | |
| | CD8$^{High}$ | V1 | 0.0034 | 0.0021 | 0.0126 |
| | | V3 | < 0.0001 | < 0.0001 | 0.0003 |
| | CD4$^{+}$ CD8$^{+}$ | V1 | | | |
| | | V3 | | | **0.0329** |
| B cells | CD3$^{-}$ CD33$^{-}$ CD19$^{+}$ | V1 | | | |
| | | V3 | | | |
| NK cells | CD3$^{-}$ CD33$^{-}$ CD19$^{-}$ | V1 | 0.0455 | | |
| | | V3 | | 0.0033 | 0.0246 |

**Table S3. Cells from infected participants are less prone to divide compared to non-infected participants following a 7-day assay**

Comparison between the non-infected (n=18-28) and infected groups (n=27-38) at V1 and at V3. Values in bold represent an increase in the expansion index of the infected group compared to the non-infected group, while plain values represent a decrease. Data were analized with Mann-Whitney two-tailed unpaired t test.

| | Cell Type | B. Group | Pep JPT | Ancestral | Omicron |
|---|---|---|---|---|---|
| T cells | $CD4^+$ Total | Non-Inf | | | |
| | | Infected | | 0.0235 | |
| | $CD4^{Low}$ | Non-Inf | | | |
| | | Infected | | | |
| | $CD4^{High}$ | Non-Inf | | | |
| | | Infected | 0.0067 | 0.0023 | 0.021 |
| | $CD8^+$ Total | Non-Inf | | | |
| | | Infected | 0.0007 | | |
| | $CD8^{Low}$ | Non-Inf | | | |
| | | Infected | | | 0.0412 |
| | $CD8^{High}$ | Non-Inf | | | |
| | | Infected | 0.0493 | 0.0094 | 0.0145 |
| | $CD4^+$ $CD8^+$ | Non-Inf | | | |
| | | Infected | | | |
| B cells | $CD3^-$ $CD33^-$ $CD19^+$ | Non-Inf | | | |
| | | Infected | | | |
| NK cells | $CD3^-$ $CD33^-$ $CD19^-$ | Non-Inf | | | |
| | | Infected | | | |

**Table S4. Cells from infected participants divide less following infection**

Comparison between both visits for the non-infected (n=18-28) and infected (n=27-38) groups separately. Plain values represent a decrease in the expansion index at V3. Data were analized with Wilcoxon two-tailed paired t test.

| Category | Variable | Condition | AUC | Std. Error | 95% C.I. | P-value |
|---|---|---|---|---|---|---|
| **Antibody detection** | IgG | Anti-RBD | 0.7889 | 0.05837 | 0.6744 - 0.9033 | < 0.0001 |
| | | Anti-S | 0.7566 | 0.06126 | 0.6365 - 0.8767 | 0.0002 |
| | | Anti-N | 0.8834 | 0.03972 | 0.8056 - 0.9613 | < 0.0001 |
| | IgG3 | Anti-N | 0.9413 | 0.02854 | 0.8854 - 0.9973 | < 0.0001 |
| **Microneutralization** | Ancestral | - | 0.7819 | 0.06008 | 0.6641 - 0.8997 | < 0.0001 |
| | Delta | - | 0.8295 | 0.05222 | 0.7272 - 0.9319 | < 0.0001 |
| | Omicron | - | 0.89 | 0.0414 | 0.8089 - 0.9712 | < 0.0001 |
| **PMN** | IL-8 | R848 1 µg/mL | 0.6379 | 0.06495 | 0.5106 - 0.7652 | 0.0451 |
| **PBMC** | TNF | R848 1 µg/mL | 0.6611 | 0.07141 | 0.5211 - 0.8010 | 0.033 |
| **Phenotype** | $CD33^+$ | Resting | 0.6793 | 0.06457 | 0.5528 - 0.8059 | 0.0092 |
| | | Ancestral | 0.6509 | 0.06644 | 0.5207 - 0.7811 | 0.0283 |
| | | Omicron | 0.6452 | 0.06635 | 0.5151 - 0.7752 | 0.0349 |
| | $CD4^+$ | Ancestral | 0.6436 | 0.06523 | 0.5158 - 0.7715 | 0.0369 |
| | $CD4^{High}$ | Ancestral | 0.6386 | 0.06601 | 0.5093 - 0.7680 | 0.044 |
| | $CD19^+$ | Resting | 0.6517 | 0.06579 | 0.5227 - 0.7806 | 0.0275 |
| | | Pep JPT | 0.6751 | 0.06626 | 0.5452 - 0.8050 | 0.0109 |
| | | Ancestral | 0.6517 | 0.06562 | 0.5231 - 0.7803 | 0.0275 |
| | | Omicron | 0.6525 | 0.06682 | 0.5215 - 0.7834 | 0.0267 |
| **Proliferation index** | $CD4^{Low}$ | Pep JPT | 0.7302 | 0.06412 | 0.6046 - 0.8559 | 0.002 |
| | $CD4^{High}$ | Pep JPT | 0.6631 | 0.06759 | 0.5306 - 0.7955 | 0.0244 |
| | | Omicron | 0.7577 | 0.06068 | 0.6388 - 0.8767 | 0.0004 |
| | $CD8^+$ | Pep JPT | 0.6565 | 0.06754 | 0.5241 - 0.7889 | 0.0308 |
| | | Ancestral | 0.6461 | 0.0695 | 0.5099 - 0.7824 | 0.0436 |
| | | Omicron | 0.6829 | 0.0656 | 0.5543 - 0.8115 | 0.0121 |
| | $CD8^{High}$ | Pep JPT | 0.7227 | 0.06443 | 0.5965 - 0.8490 | 0.0021 |
| | | Ancestral | 0.7063 | 0.06407 | 0.5807 - 0.8319 | 0.0044 |
| | | Omicron | 0.6984 | 0.06519 | 0.5706 - 0.8261 | 0.0065 |
| | NK | Ancestral | 0.6476 | 0.07099 | 0.5084 - 0.7867 | 0.0417 |
| **ELISA proliferation** | TNF | Pep JPT | 0.6576 | 0.06609 | 0.5280 - 0.7871 | 0.022 |
| | | Ancestral | 0.6628 | 0.06384 | 0.5376 - 0.7879 | 0.0169 |
| | | Omicron | 0.6569 | 0.06411 | 0.5312 - 0.7825 | 0.0213 |
| | IFNγ | Pep JPT | 0.7318 | 0.06239 | 0.6095 - 0.8541 | 0.0008 |
| | | Ancestral | 0.7815 | 0.05651 | 0.6708 - 0.8923 | < 0.0001 |
| | | Omicron | 0.7764 | 0.05519 | 0.6682 - 0.8846 | < 0.0001 |

**Table S5. Several immune parameters discriminate between infected and non-infected individuals at V3**

Receiver operating characteristic analysis performed with GraphPad Prism 10.5.0 with confidence interval calculated with the Wilson/Brown method. AUC: area under the curve.

| Assay | Oligonucleotide | Sequence | Final Concentration (nM) |
|---|---|---|---|
| E gene | E_Sarbeco_F1 (2) | 5' ACAGGTACGTTAATAGTTAATAGCGT 3' | 800 |
| | E_IDT_R | 5' ACAATCGAAGCGCAGTAAGG 3' | 800 |
| | E_IDT_P | 5' TGCTTTCGTGGTATTCTTGCTAGTTACACT 3' (FAM-ZEN-IABQ) | 200 |
| N gene | N_IDT_F | 5' CGTACTGCCACTAAAGCATACA 3' | 400 |
| | N_IDT_R | 5' GCGGCCAATGTTTGTAATCAG3' | 400 |
| | N_IDT_P | 5' AGACGTGGTCCAGAACAAACCCAA 3' (HEX-ZEN-IABQ) | 200 |

**Table S6. Primers and probes for SARS-CoV-2 quantification**

| Antibody | Fluorochrome | Source | Identifier |
|---|---|---|---|
| Mouse monoclonal anti-human CD33 | PE | Invitrogen | Cat# 12-0338-42<br>RRID: AB_10855036<br>Clone WM-53<br>Lot: 2400600 |
| Mouse monoclonal anti-human CD3 | PerCP-Cy5.5 | Invitrogen | Cat# 45-0036-42<br>RRID: AB_1518742<br>Clone SK7<br>Lot: 2407521 |
| Mouse monoclonal anti-human CD4 | PE-Cy7 | Invitrogen | Cat# 25-0049-42<br>RRID: AB_1659695<br>Clone RPA-T4<br>Lot: 239648 |
| Mouse monoclonal anti-human CD8a | APC | Invitrogen | Cat# 17-0086-42<br>RRID: AB_10667892<br>Clone OKT8<br>Lot: 2410239 |
| Mouse monoclonal anti-human CD19 | APC-eFluor780 | Invitrogen | Cat# 47-0198-42<br>RRID: AB_10719114<br>Clone SJ25C1<br>Lot: 2427148 |
| Mouse IgG1 κ isotype control | PerCP-Cy5.5 | Invitrogen | Cat#45-4714-82<br>Clone P3.6.2.8.1<br>Lot: 2011175 |
| Mouse IgG1 κ isotype control | PE-Cy7 | Invitrogen | Cat#25-4714-80<br>Clone P3.6.2.8.1<br>Lot: 2298238 |
| Mouse IgG2 κ isotype control | APC | Invitrogen | Cat#17-4724-81<br>Clone eBM2a<br>Lot: 2045450 |
| Mouse IgG1 κ isotype control | APC-eFluor 780 | Invitrogen | Cat#47-4714-82<br>Clone P3.6.2.8.1<br>Lot: 2062716 |
| Mouse IgG1 κ isotype control | PE | Invitrogen | Cat#12-4714-82<br>Clone P3.6.2.8.1<br>Lot: 2356239 |

**Table S7. Flow cytometry antibody panel and isotype controls**

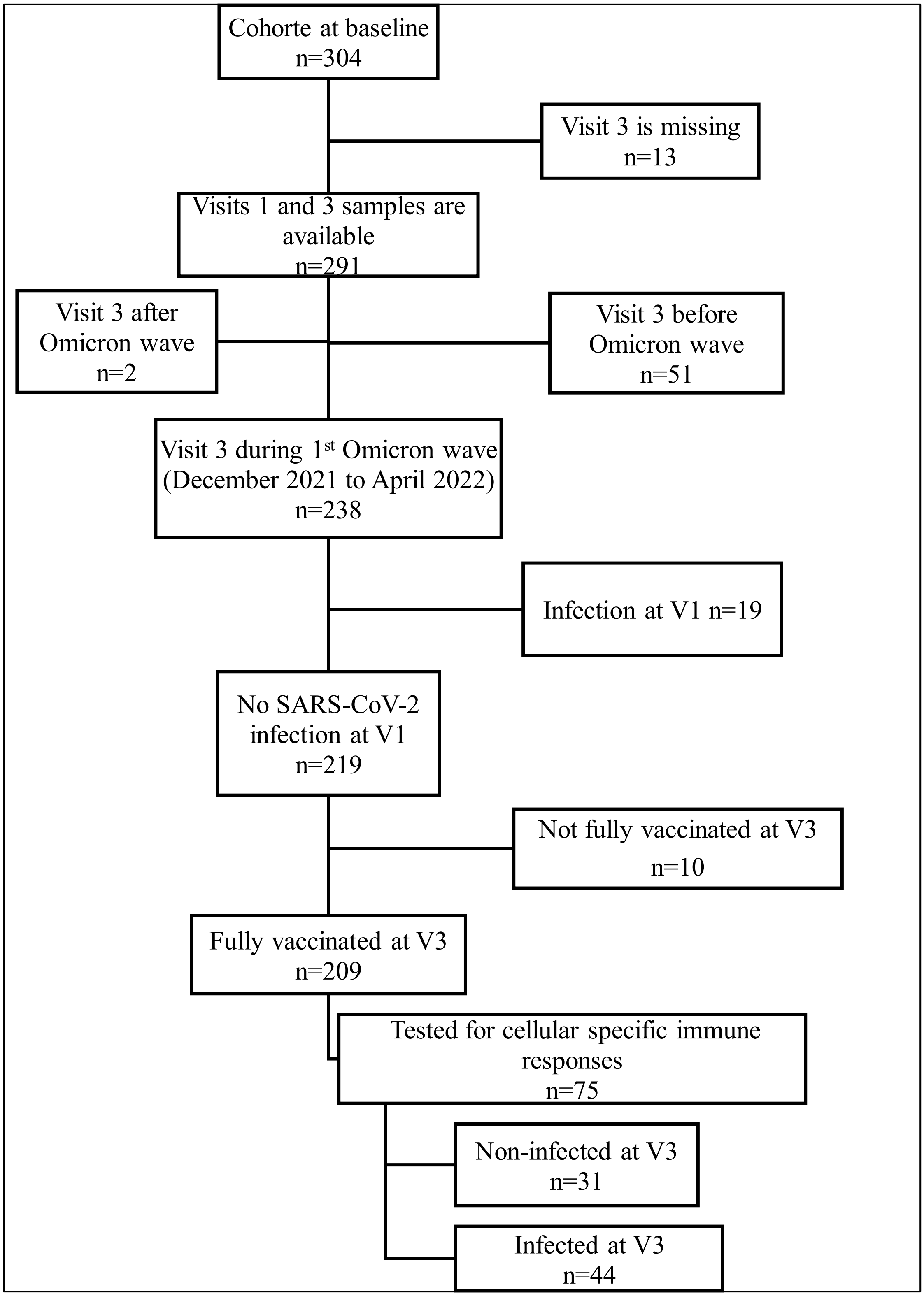


**Figure S1. Participant selection flow chart**

From the 304 participants of the cohort at V1, 75 were selected based on the availability of samples, date of visits, vaccination and infection status.

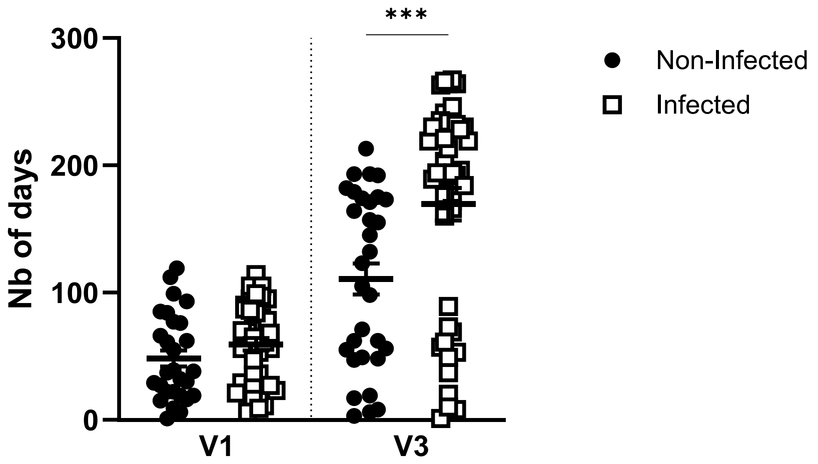


**Figure S2. Infected participants had a longer elapsed time since their last vaccine dose at V3 compared to non-infected**

Time elapsed between the last vaccine dose received and the visits for non-infected (full dots; n=28-31) and infected (empty squares; n=42-44) participants. Each dot represents a participant. Data are shown as mean ± SEM, and were analyzed with Mann-Whitney two-tailed unpaired t test. *** $p < 0.001$.

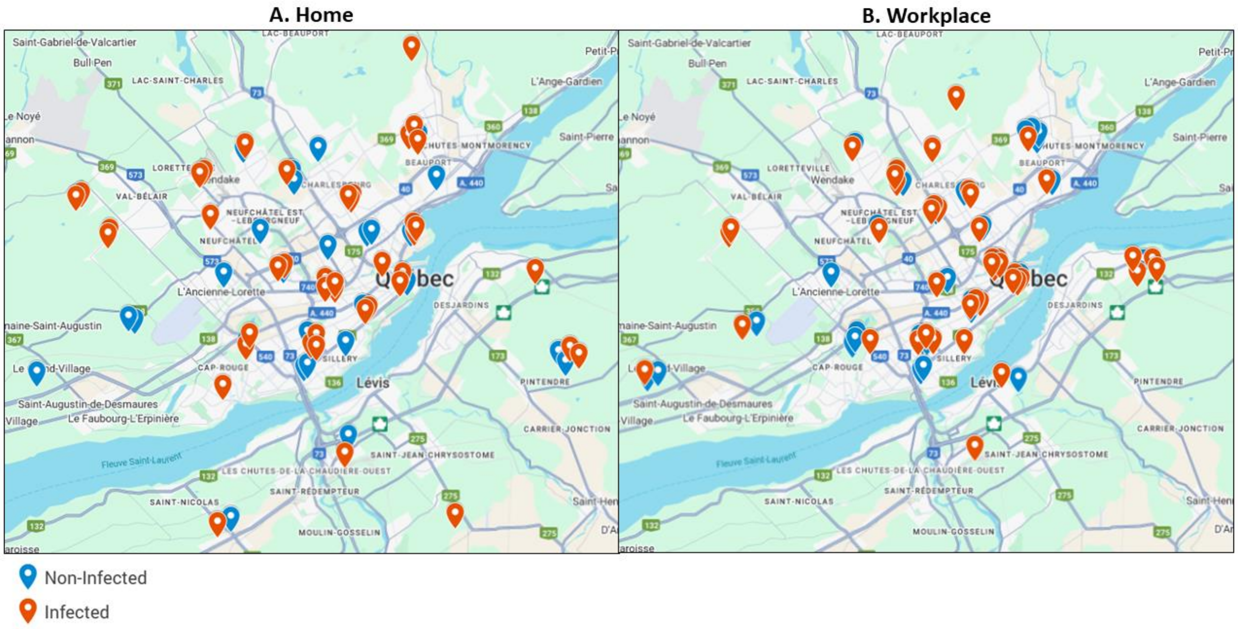


**Figure S3. Localization of participant's home and workplace zip codes**

The three first digits of subjects' home and workplace zip codes were mapped using Google My Maps.

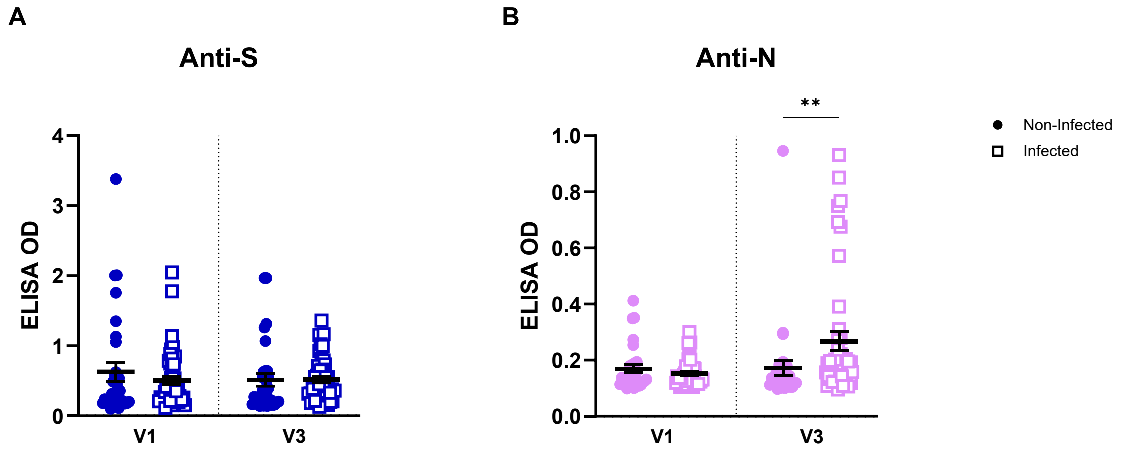


**Figure S4. Infected individuals have higher levels of anti-N circulating antibodies**

Serum antibody levels in vaccinated non-infected (full dots; n=31) and infected (empty squares; n=44) participants before (V1) and during (V3) Omicron predominance by the in-house ELISA method. **A)** Anti-spike (S; ancestral) IgG levels. **B)** Anti-nucleocapsid (N) IgG levels. Each dot represents a participant. Data were analized with Mann-Whitney two-tailed unpaired t test. ** $p < 0.01$. OD: optical density.

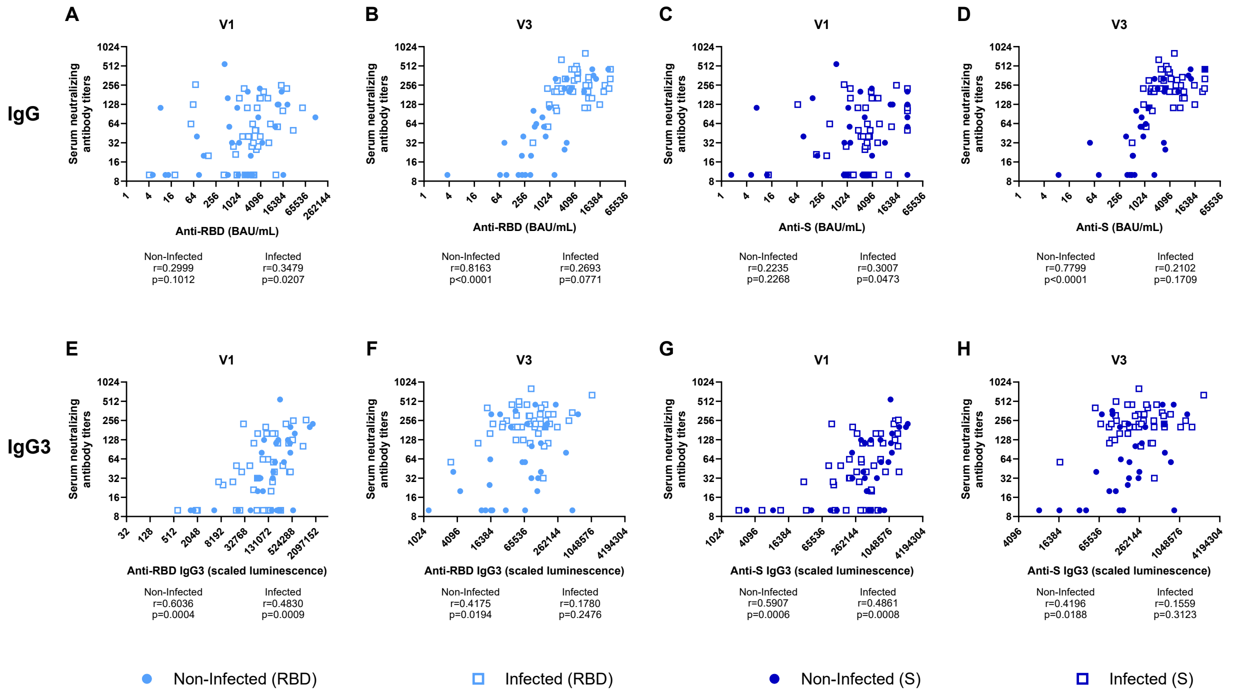


**Figure S5. Protective immunity is associated with preserved coupling between Spike recognition and neutralizing activity**

Correlations between the microneutralization results against the ancestral virus (y axis) and the serum antibody levels (x axis) of **A)** anti-RBD at V1, **B)** anti-RBD at V3, **C)** anti-S at V1, and **D)** anti-S at V3 for non-infected (n=44) and infected (n=31) groups. Similarly, correlations were done with microneutralization results against the ancestral virus and the IgG3 antibody levels for **E)** anti-RBD at V1, **F)** anti-RBD at V3, **G)** anti-S at V1, and **H)** anti-S at V3. Each dot represents a participant for non-infected (n=44) and infected (n=30-31) groups. Data were analized with Spearman correlation. P-value < 0.05 is significant. BAU: binding antibody units.

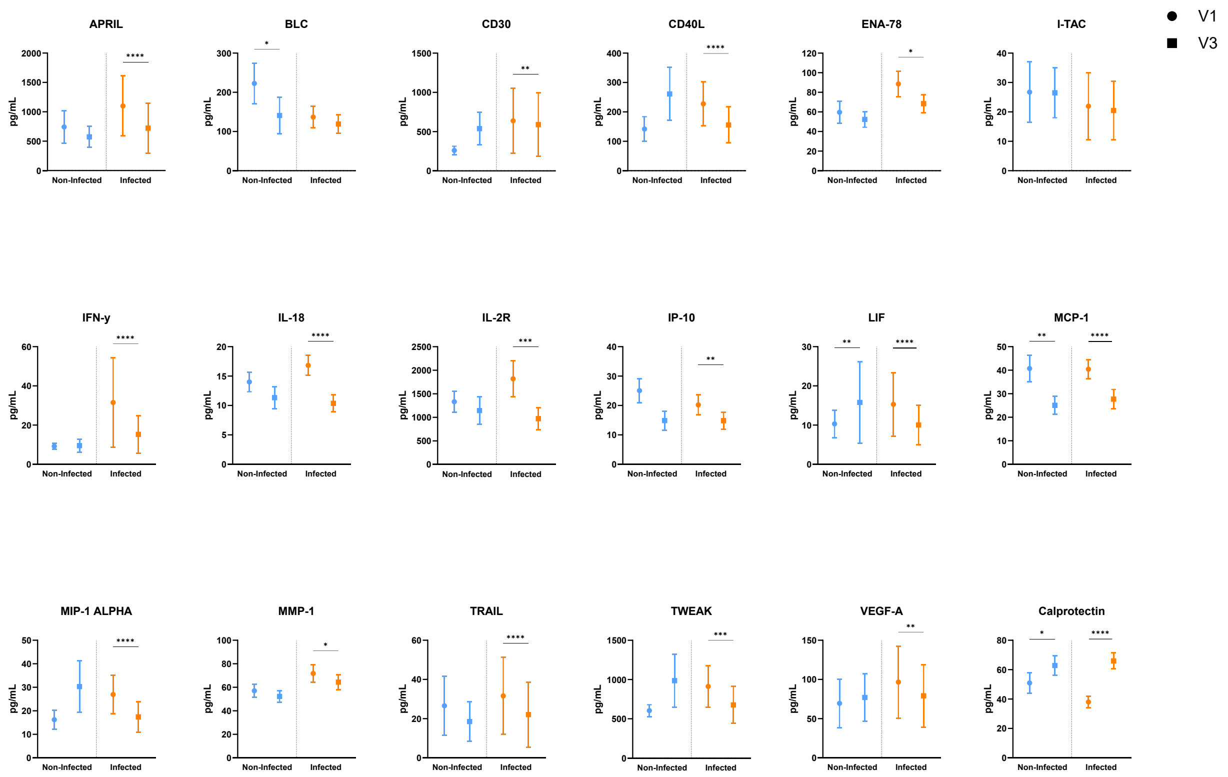


**Figure S6. Infected individuals show a greater number of downregulated cytokines at V3**

Mean plasma cytokine levels in non-infected (blue, n=25) and infected (green, n=40) individuals, before (V1) and during Omicron predominance (V3). Data are shown as mean ± SEM. Wilcoxon test was performed. * $p < 0.05$; ** $p < 0.01$; *** $p < 0.001$; **** $p < 0.0001$.

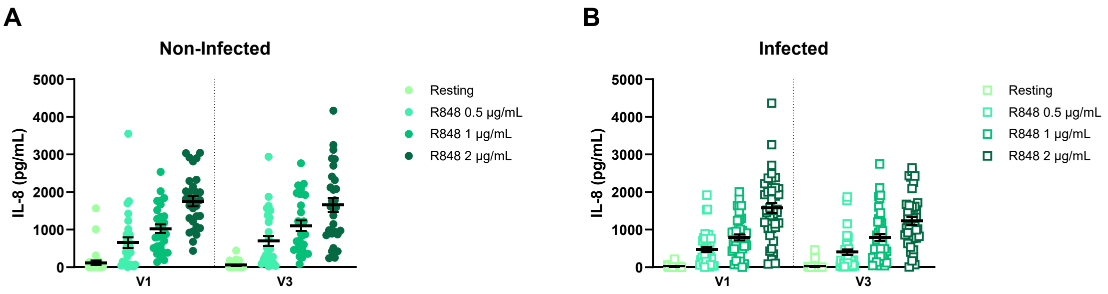


**Figure S7. PMN IL-8 production remained stable between V1 and V3 in both study groups**

Comparison of IL-8 levels produced by PMNs at V1 and V3 after 24h of stimulation with different concentrations of R848. **A)** IL-8 measures from non-infected participants (n=29). **B)** IL-8 measures from infected participants (n=42). Each dot represents a participant. Data are shown as mean ± SEM and were analyzed using Mann-Whitney two-tailed unpaired t test.

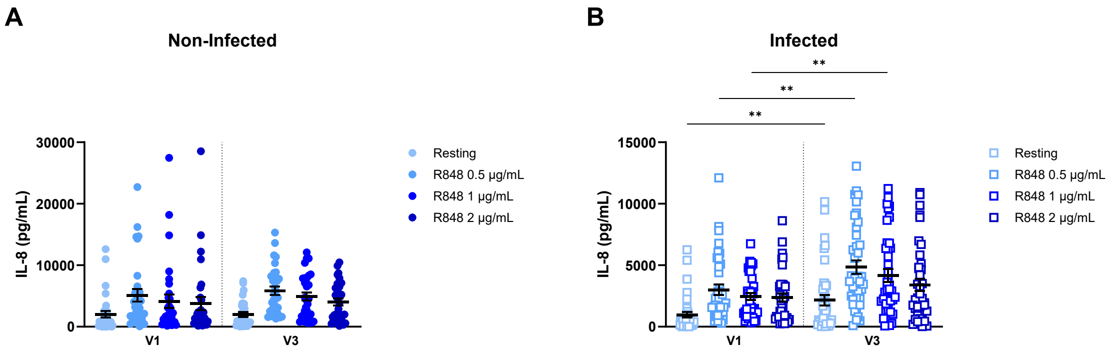


**Figure S8. PBMC IL-8 production increases after infection**

Comparison of IL-8 levels produced by PBMCs at V1 and V3 after 24h of stimulation with different concentrations of R848. **A)** IL-8 measures from non-infected participants (n=31). **B)** IL-8 measures from infected participants (n=39). Each dot represents a participant. Data are shown as mean ± SEM and were analyzed using Mann-Whitney two-tailed unpaired t test. ** $p < 0.01$.

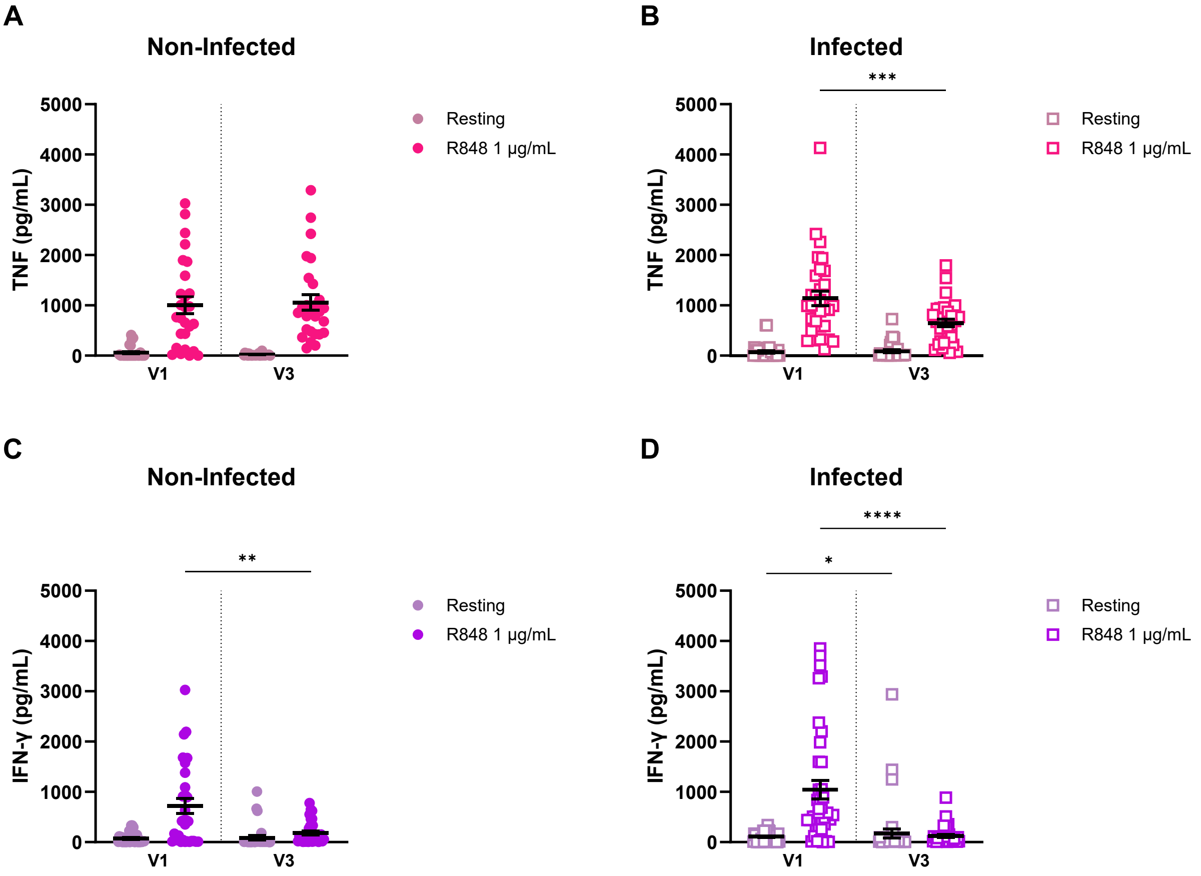


**Figure S9. Decreased PBMC TNF and IFN-γ production at V3**

Measures of TNF and IFN-γ secreted by PBMCs following 24h in resting or R848 stimulation conditions for non-infected and infected groups. **A)** TNF concentrations for non-infected subjects at V1 and V3 (n=27). **B)** TNF concentrations for infected subjects at V1 and V3 (n=32). **C)** IFN-γ concentrations for non-infected subjects at V1 and V3 (n=31). **D)** IFN-γ concentrations for infected subjects at V1 and V3 (n=38). Each dot represents a participant. Data were analized with Mann-Whitney two-tailed unpaired t test. * $p < 0.05$; ** $p < 0.01$; *** $p < 0.001$; **** $p < 0.0001$.

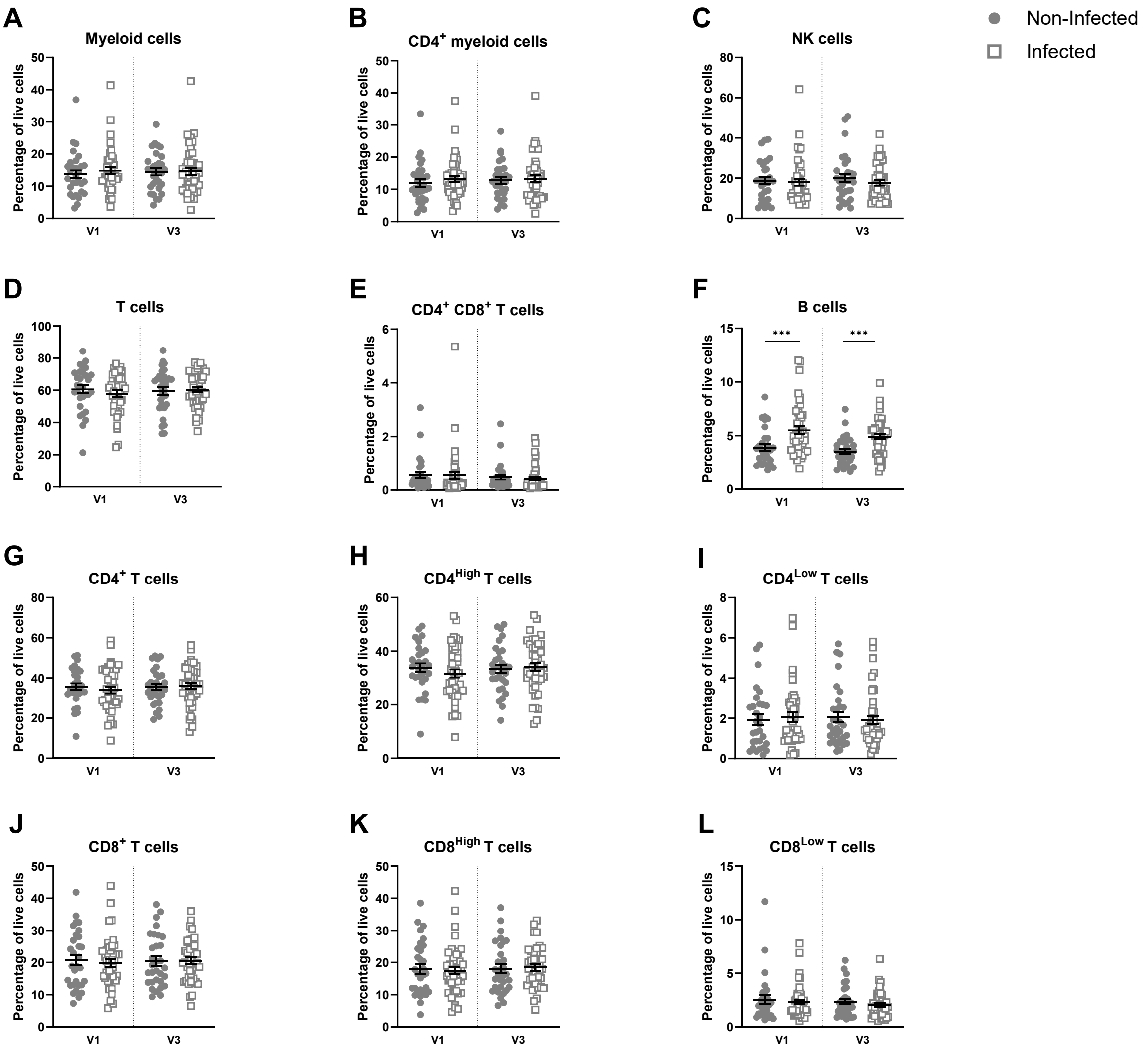


**Figure S10. Infected individuals maintain a higher percentage of B cells before and after infection**

Phenotypes of PBMCs at the start of the proliferation assay, before stimulation. Each phenotype is shown as the percentage of live cells per individual. Proportion of **A)** myeloid cells ($CD33^+$); **B)** $CD4^+$ myeloid cells ($CD33^+$ $CD4^+$); **C)** NK cells ($CD33^-$ $CD3^-$ $CD19^-$); **D)** T cells ($CD33^-$ $CD3^+$); **E)** double positive T cells ($CD33^-$ $CD3^+$ $CD4^+$ $CD8^+$); **F)** B cells ($CD33^-$ $CD3^-$ $CD19^+$); **G)** $CD4^+$ T cells ($CD33^-$ $CD3^+$ $CD4^+$); **H)** $CD4^{High}$ T cells ($CD33^-$ $CD3^+$ $CD4^{High}$); **I)** $CD4^{Low}$ T cells ($CD33^-$ $CD3^+$ $CD4^{Low}$); **J)** $CD8^+$ T cells ($CD33^-$ $CD3^+$ $CD8^+$); **K)** $CD8^{High}$ T cells ($CD33^-$ $CD3^+$ $CD8^{High}$); **L)** $CD8^{Low}$ T cells ($CD33^-$ $CD3^+$ $CD8^{Low}$). Non-infected: V1 n=30, V3 n=31; infected V1 n=44, V3: n=42. Each dot represents a participant. Data were analyzed with Mann-Whitney two-tailed unpaired t test. *** $p < 0.001$.

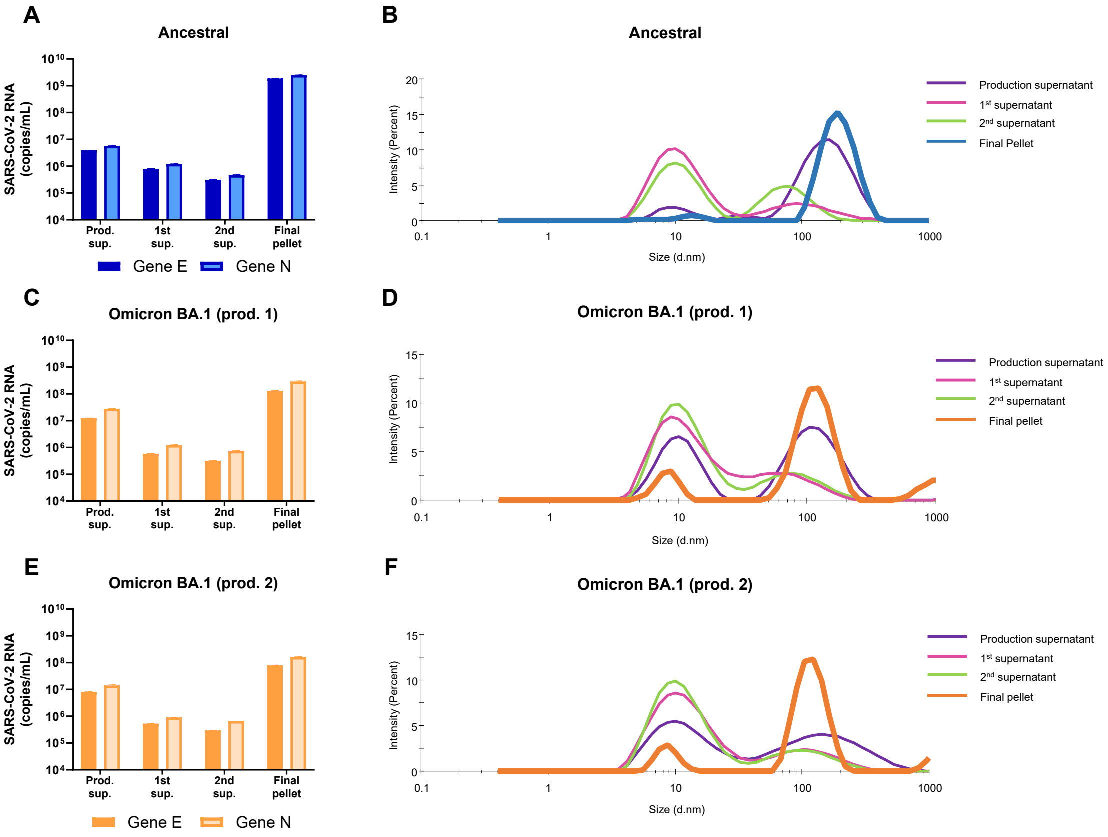


**Figure S11. Characterization of the inactivated SARS-CoV-2 preparations used for the proliferation assay**

SARS-CoV-2 RNA was quantified by RT-qPCR in the production supernatant, the supernatant of the first and second ultracentrifugation, and the final pellet to assess the virus purification efficiency. One ancestral SARS-CoV-2 (**A**), and two Omicron BA.1 preparations (**C**, **E**) were used. Viral samples were also analyzed by dynamic light scattering (DLS) for the ancestral virus (**B**) and both productions of the Omicron BA.1 variant (**D**, **F**).

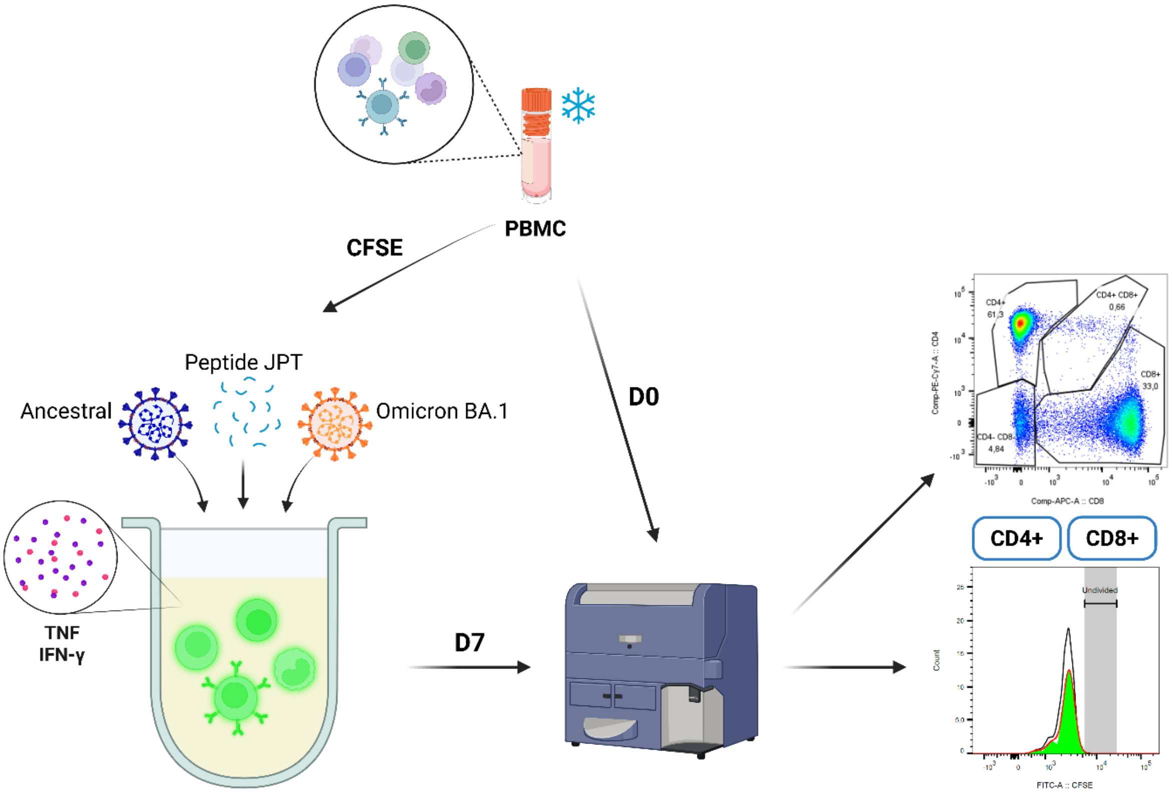


**Figure S12. Adaptive immunity experimental setup**

Schematic representation of the experimental setup to study the adaptive immunity. Frozen PBMCs were thawed, a fraction was kept as a day 0 (D0) as control, and the rest was stained with CFSE and incubated for seven days in four different conditions (resting, pep JPT, inactivated ancestral or Omicron BA.1 viruses). At day 7 (D7), cells were stained for flow cytometry to look at proliferation and phenotypes (myeloid cells: $CD33^+$; $CD4^+$ myeloid cells: $CD33^+$ $CD4^+$; $CD4^+$ T cells: $CD33^-$ $CD3^+$ $CD4^+$; $CD8^+$ T cells: $CD33^-$ $CD3^+$ $CD8^+$; $CD4^+$ $CD8^+$ T cells: $CD33^-$ $CD3^+$ $CD4^+$ $CD8^+$; B cells: $CD33^-$ $CD3^-$ $CD19^+$; NK cells: $CD33^-$ $CD3^-$ $CD19^-$). The culture supernatant was also collected to measure TNF and IFN-γ production by ELISA.

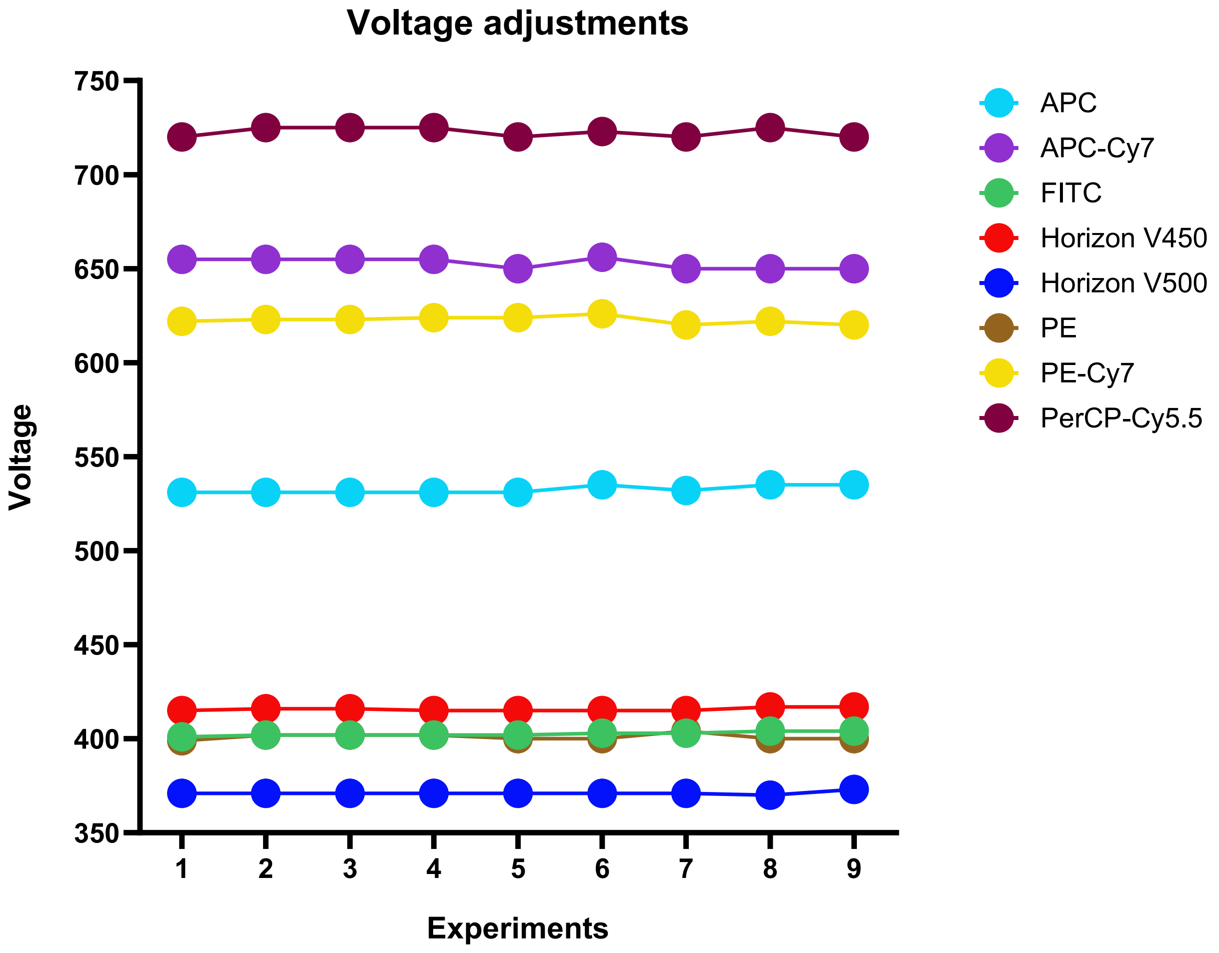


**Figure S13. Voltage adjustments for all cytometry experiments**

Voltages used throughout the cytometry experiments following rainbow beads calibration control.

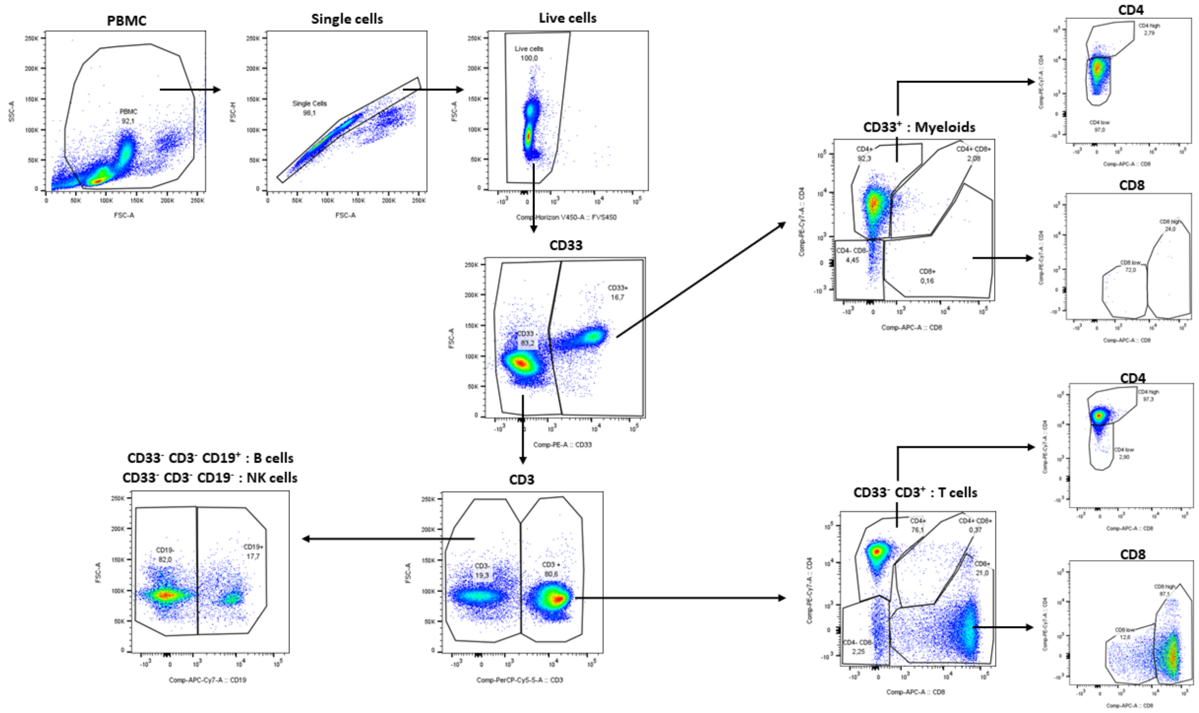


**Figure S14. Gating strategy**

PBMCs are first selected, avoiding debris, followed by single cells and live cells. $CD33^+$ cells are selected to obtain myeloid cells. From the $CD33^-$ cells, $CD3^+$ cells are selected to obtain T cells, and from those, $CD4^+$ and $CD8^+$ are separated. They are further gated to obtain high and low expression of both markers. From the $CD3^-$ cells, CD19 marker is used to differentiate between B cells ($CD19^+$) and NK cells ($CD19^-$).
